# COVID-19 Variant Detection with a High-Fidelity CRISPR-Cas12 Enzyme

**DOI:** 10.1101/2021.11.29.21267041

**Authors:** Clare L. Fasching, Venice Servellita, Bridget McKay, Vaishnavi Nagesh, James P. Broughton, Alicia Sotomayor-Gonzalez, Baolin Wang, Noah Brazer, Kevin Reyes, Jessica Streithorst, Rachel N. Deraney, Emma Stanfield, Carley G. Hendriks, Steve Miller, Jesus Ching, Janice S. Chen, Charles Y. Chiu

## Abstract

Laboratory tests for the accurate and rapid identification of SARS-CoV-2 variants can potentially guide the treatment of COVID-19 patients and inform infection control and public health surveillance efforts. Here we present the development and validation of a rapid COVID-19 variant DETECTR^®^ assay incorporating loop-mediated isothermal amplification (LAMP) followed by CRISPR-Cas12 based identification of single nucleotide polymorphism (SNP) mutations in the SARS-CoV-2 spike (S) gene. This assay targets the L452R, E484K/Q/A, and N501Y mutations that are associated with nearly all circulating viral lineages and identifies the two circulating variants of concern, Delta and Omicron. In a comparison of three different Cas12 enzymes, only the newly identified enzyme CasDx1 was able to accurately identify all targeted SNP mutations. An analysis pipeline for CRISPR-based SNP identification from 139 clinical samples yielded an overall SNP concordance of 98% and agreement with SARS-CoV-2 lineage classification of 138/139 compared to viral whole-genome sequencing. We also showed that detection of the single E484A mutation was necessary and sufficient to accurately identify Omicron from other major circulating variants in patient samples. These findings demonstrate the utility of CRISPR-based DETECTR^®^ as a faster and simpler diagnostic than sequencing for SARS-CoV-2 variant identification in clinical and public health laboratories.

## Introduction

The emergence of new SARS-CoV-2 variants threatens to substantially prolong the COVID-19 pandemic. SARS-CoV-2 variants, especially Variants of Concern (VOCs)^1, 2^, have caused resurgent COVID-19 outbreaks in the United States^2–5^ and worldwide^1, 6, 7^, even in populations with a high proportion of vaccinated individuals^8–11^. Mutations in the spike protein, which binds to the human ACE2 receptor, can render the virus more infectious and/or more resistant to antibody neutralization, resulting in increased transmissibility^12^, and/or escape from immunity, whether vaccine-mediated or naturally acquired immunity^13, 14^. Variant identification can also be clinically significant, as some mutations substantially reduce the effectiveness of available monoclonal antibody therapies for the disease^15^.

Tracking the evolution and spread of SARS-CoV-2 variants in the community can inform public policy regarding testing and vaccination, as well as guide contact tracing and containment effects during local outbreaks^16, 17^. Virus whole-genome sequencing (WGS) and single nucleotide polymorphism (SNP) genotyping are commonly used to identify variants^16, 18^, but can be limited by long turnaround times and/or the requirement for bulky and expensive laboratory instrumentation. Diagnostic assays based on clustered interspaced short palindromic repeats (CRISPR)^19^ have been developed for rapid detection of SARS-CoV-2 in clinical samples^13, 20–23^, and a few have obtained Emergency Use Authorization (EUA) by the US Food and Drug Administration (FDA)^24–26^. Some advantages of these assays for use in laboratory and point of care settings include low cost, minimal instrumentation, and a sample-to-answer turnaround time of under 2 hours^20, 23, 27–29^.

Here we present the development of a CRISPR-based COVID-19 variant DETECTR^®^ assay (henceforth abbreviated as DETECTR^®^ assay) for the detection of SARS-CoV-2 mutations and evaluate its performance on a total of 139 patient respiratory swab samples using WGS as a comparator method (Fig. 1a). The assay combines RT-LAMP pre-amplification followed by fluorescent detection using a CRISPR-Cas12 enzyme. We perform a comparative evaluation of multiple candidate Cas12 enzymes and demonstrate that robust assay performance depends on the specificity of the newly identified CRISPR-Cas12 enzyme called CasDx1 in identifying key SNP mutations of functional relevance in the spike protein at amino acid positions 452, 484 and 501^30^.

**Fig. 1.**
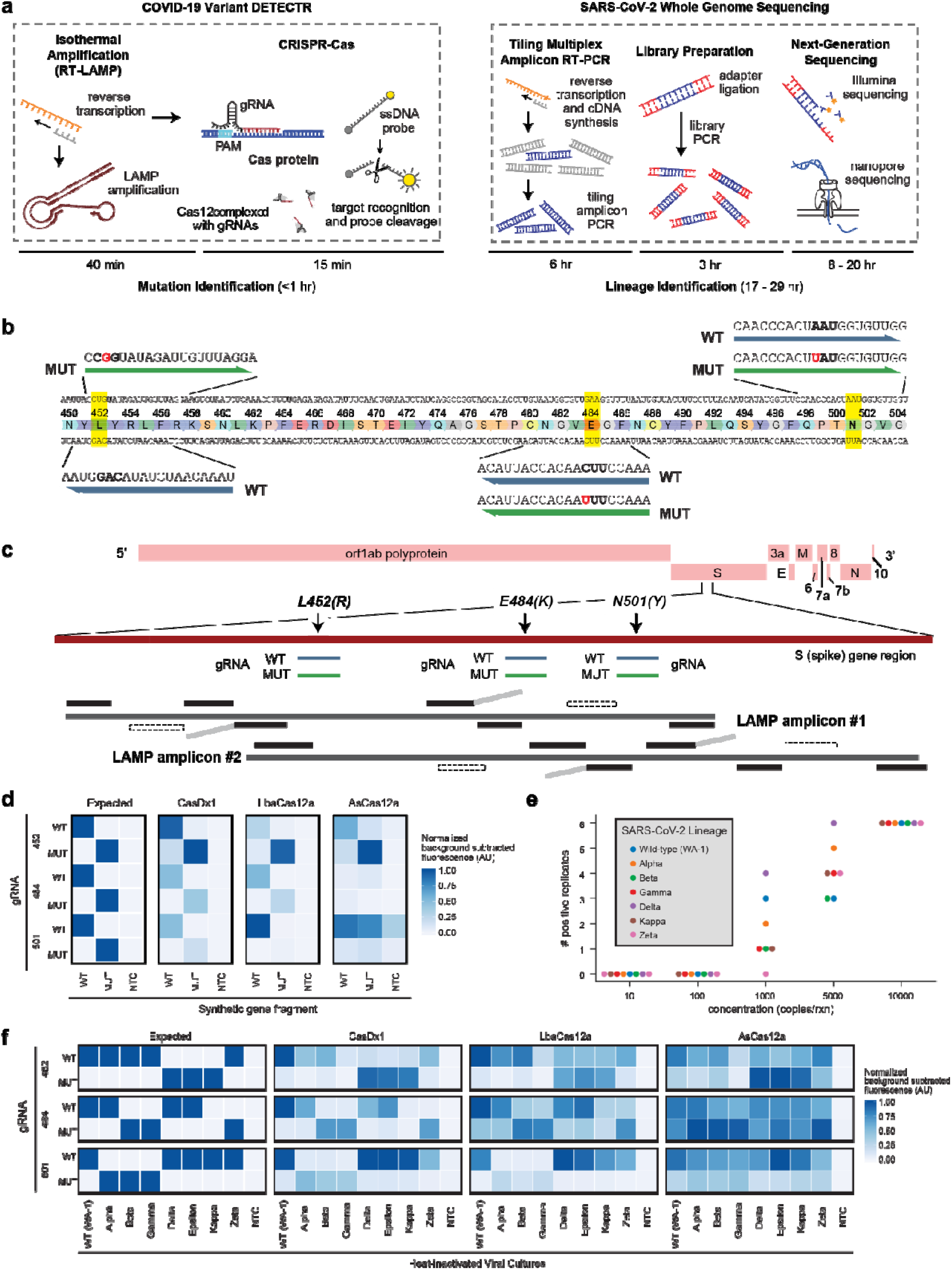
Design and Workflow for the DETECTR^®^ assay. **a**, Workflow comparison between the DETECTR^®^ assay and SARS-CoV-2 whole-genome sequencing (WGS). **b**, Schematic of CRISPR-Cas gRNA design for SARS-CoV-2 S gene mutations. **c**, Schematic of multiplexed RT-LAMP primer design showing the SARS-CoV-2 S gene mutations and gRNA positions. **d**, Heat map comparison of three different Cas12 enzymes tested using 10 nM PCR-amplified synthetic gene fragments (t = 30 minutes). **e**, Dot plot showing the number (n = 6) of positive replicates across a 4-log dynamic range of the RT-LAMP products. **f**, Heat map comparison of end-point fluorescence (t = 30 minutes) of three different Cas12 enzymes tested against heat-inactivated viral cultures. Replicates (n = 6) generated using RT-LAMP were pooled and CRISPR-Cas12 reactions were then run in triplicate (n = 3).

## Results

### Identifying the optimal CRISPR-Cas12 enzyme for SNP detection

To determine the optimal Cas12 enzyme for SNP detection, we evaluated three different CRISPR-Cas effectors with trans-cutting activity: LbCas12a, AsCas12a, and a novel Cas12 enzyme called CasDx1. We initially screened guide RNAs (gRNAs) with CasDx1 and LbCas12a for activity on synthetic gene fragments encoding regions of the SARS-CoV-2 S-gene with either wild-type (WT) or mutant (MUT) sequences at amino acid positions 452, 484, and 501 (Fig. 1b-c). From this initial activity screen, we identified the top-performing gRNAs for each S-gene variant encoding either L452R, E484K or N501Y (Fig. 1d). Further evaluation of these guides using CasDx1, LbCas12a and AsCas12a with their cognate gRNAs on synthetic gene fragments revealed differences in SNP differentiation capabilities, with CasDx1 showing the clearest SNP differentiation between wild-type (WT) and mutant (MUT) sequences for all targeted S-gene variants (Fig. 1d **and** Extended Data Fig. 1a). In comparison, LbCas12a could differentiate SNPs at positions 452 and 484, but not 501, whereas AsCas12a could only differentiate the SNP at position 452 (Fig. 1d **and** Extended Data Fig. 1a).

We next tested SNP differentiation capabilities on heat-inactivated viral cultures using the full DETECTR^®^ assay, consisting of RNA extraction, multiplexed RT-LAMP amplification (Fig. 1c), and CRISPR-Cas12 detection with guide RNAs targeting part of the spike receptor-binding domain (RBD) (Fig. 1b). The LAMP primer design incorporated two sets of six primers each, with both sets generating overlapping spike RBD amplicons that spanned the L452R, E484K, and N501Y mutations. We chose to adopt a redundant LAMP design for two reasons: first, this approach was shown to improve detection sensitivity in initial experiments; second, we sought to increase assay robustness given the continual emergence of escape mutations in the spike RBD throughout the course of the pandemic^13^. The tested viral cultures included an ancestral SARS-CoV-2 lineage (WA-1) containing the wild-type spike protein (D614) targeted by the approved mRNA (BNT162b2 from Pfizer or mRNA-1273 from Moderna)^31, 32^ and DNA adenovirus vector (Ad26.COV2.S from Johnson and Johnson)^33^ vaccines, variants being monitored (VBMs) that were previously classified as VOCs or variants of interest (VOIs), including Alpha (B.1.1.7), Beta (B.1.351), Gamma (P.1), Epsilon (B.1.427 and B.1.429), Kappa (B.1.617.1), and Zeta (P.2) lineages, and the current VOC Delta (B.1.617.2) lineage^34^. Heat-inactivated viral culture samples representing the seven SARS-CoV-2 lineages were quantified by digital droplet PCR across a 4-log dynamic range and used to evaluate the analytical sensitivity of the pre-amplification step. RT-LAMP amplification was evaluated using six replicates from each viral culture. We observed consistent amplification for all seven SARS-CoV-2 lineages with 10,000 copies of target input per reaction (200,000 copies/mL) (Fig. 1e), which is comparable to the target input of >200,000 copies/mL viruses (<30 Ct value) required for sequencing workflows used in SARS-CoV-2 variant surveillance^35, 36^.

To evaluate the specificity of the different Cas12 enzymes, amplified material from each viral culture was pooled and the SNPs resulting in the L452R, E484K and N501Y mutations were detected using CasDx1, LbCa12a and AsCas12a. Similar to the results found using gene fragments, CasDx1 correctly identified the wild-type (WT) and mutational (MUT) targets at positions 452, 484 and 501 in each LAMP-amplified, heat-inactivated viral culture (Fig. 1f **and** Extended Data Fig. 1b). In comparison, LbCas12a could differentiate WT from MUT at position 501 on LAMP-amplified viral cultures but showed much higher background for the WT target at position 452 and higher background for both WT and MUT targets at position 484 for (Fig. 1f **and** Extended Data Fig. 1b). Additionally, AsCas12a could differentiate WT from MUT targets at position 452 albeit with substantial background but was unable to differentiate WT from MUT targets at positions 484 and 501 (Fig. 1f **and** Extended Data Fig. 1b). From these data, we concluded that CasDx1 would provide more consistent and accurate calls for the L452R, E484K and N501Y mutations. We thus proceeded to further develop the assay using only the high-fidelity CasDx1 enzyme.

### Data analysis pipeline for calling COVID-19 variant SNPs with the DETECTR^®^ assay

To develop a data analysis pipeline for calling SARS-CoV-2 SNP mutations and assign lineage classifications with the DETECTR^®^ assay (Fig. 2a-b), we first used data collected from SNP synthetic gene fragment controls (n = 279) that included all mutational combinations of 452, 484 and 501 (**see Methods**). Based on the control sample data, we generated allele discrimination plots^37, 38^ to define boundaries that separated the WT and MUT signals (Extended Data Fig. 4a). Clear differentiation between WT and MUT signals was observed when plotting the ratio against the average of the WT and MUT transformed values on a mean average (MA) plot^37, 38^ (Extended Data Fig. 4b), with 100% concordance for SNP identity at positions 452, 484, and 501 for the control samples.

**Fig. 2.**
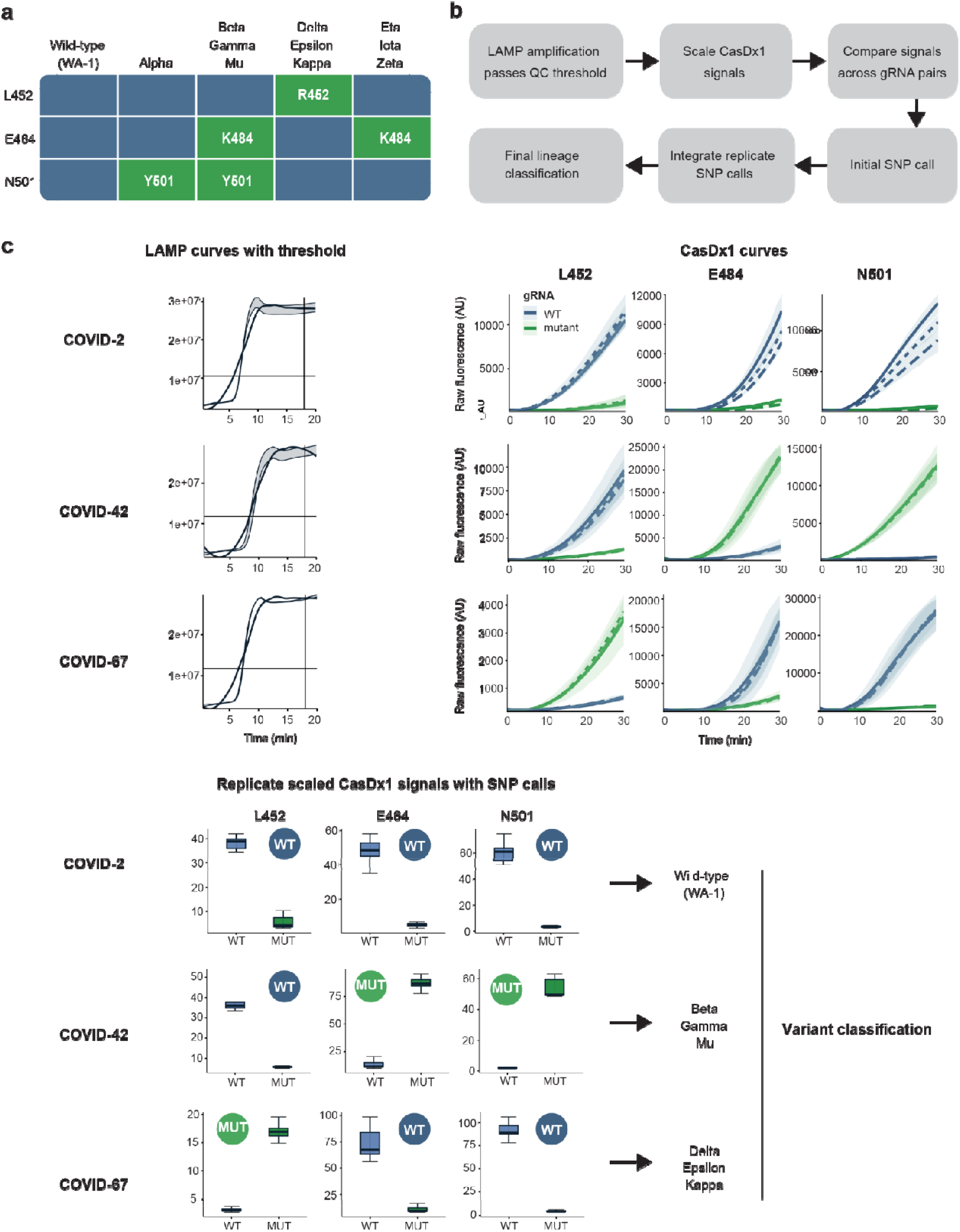
DETECTR^®^ data analysis pipeline for SARS-CoV-2 SNP mutation calling. **a**, Interpretation table summarizing the SARS-CoV-2 mutations in this study associated with the corresponding lineage classification. **b**, Schematic of data analysis pipeline describing the RT-LAMP QC and subsequent CasDx1 signal scaling. The scaled signals were compared across SNPs and the calls were made for each RT-LAMP replicate. The combined replicate calls defined the mutation call, which informed the final lineage classification. **c**, Three representative clinical samples of different SARS-CoV-2 lineages depict the workflow of the DETECTR^®^ assay. Raw fluorescence curves of each sample run in RT-LAMP amplification and subsequent triplicate DETECTR^®^ reactions targeting both WT and MUT SNPs for L452(R), E484(K), and N501(Y). Box plot visualization of the end point fluorescence in DETECTR^®^ across each SNP for the three representative clinical samples. Calls were made for each SNP by evaluating the median values of the DETECTR^®^ calls and overall calls through the LAMP replicates, and given a designation of WT, MUT, or NoCall. Final calls are made on the lineage determined by each SNP. Blue represents WT and green represents MUT, with RT-LAMP replicates (n = 3), CasDx1 replicates (n = 3 per LAMP replicate) and shading around kinetic curves indicates ±1.0 SD.

### Performance evaluation of the DETECTR^®^ assay using clinical samples

Next, we assembled a blinded dataset consisting of 93 COVID-19 positive clinical samples (previously analyzed by viral WGS) and the SNP controls run in parallel. These samples were extracted, amplified in triplicate RT-LAMP reactions (Extended Data Fig. 2), and processed further as triplicate CasDx1 reactions for each LAMP replicate (Extended Data Fig. 3). A total of nine replicates were thus generated for each sample to detect WT or MUT SNPs at positions 452, 484, and 501. The DETECTR^®^ data analysis pipeline was then applied to each sample to provide a final lineage categorization (Fig. 2a-c). For a biological RT-LAMP replicate to be designated as either WT or MUT, the same call needed to be made from all three technical CasDx1 replicates (Extended Data Fig. 5a). A final SNP mutation call was made based on ≥1 of the same calls from the three biological replicates, with replicates that were designated as a No Call ignored (Extended Data Fig. 5a-c). After excluding two samples that were considered invalid because the fluorescence intensity from RT-LAMP amplification did not reach a pre-established threshold determined using receiver-operator characteristic (ROC) curve analysis (Extended Data Fig. 2 and Extended Data Fig. 6), we evaluated a total of 807 CasDx1 signals from the 91 remaining clinical samples, generating up to 9 replicates for each clinical sample (Extended Data Fig. 5b). Differentiation of WT and MUT signals according to the allele discrimination plots was more pronounced at positions 484 and 501 than position 452 (Extended Data Fig. 4), whereas the MA plots, generated by transforming the data onto M (log ratio) and A (mean average) scales, showed clear separation of WT and MUT calls for all three positions (Fig. 3a **and** Extended Data Fig. 4). The variant calls made on each sample were consistent with the difference in median values of the log-transformed signals as determined using the data analysis pipeline (Extended Data Fig. 7).

**Fig. 3.**
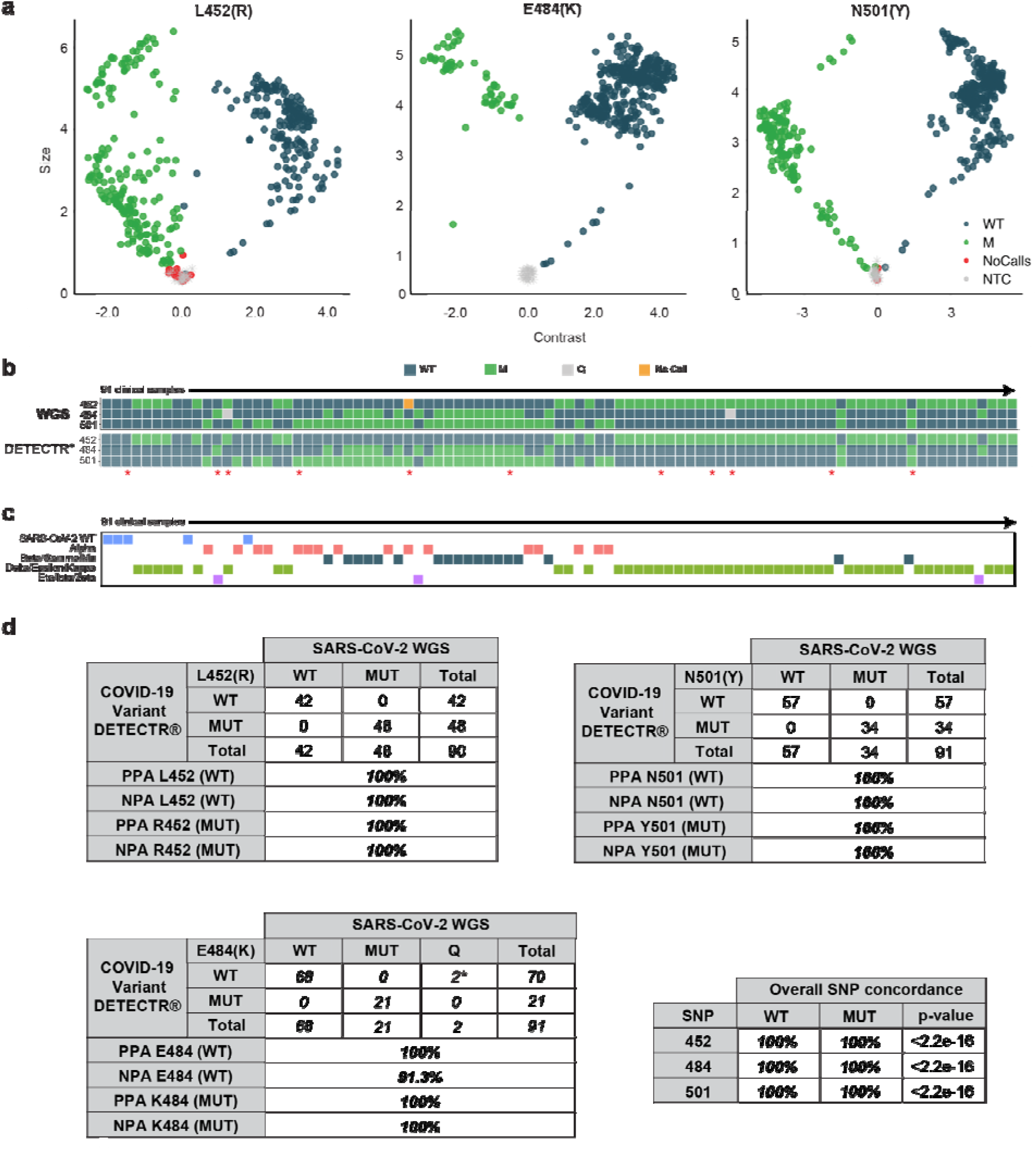
Comparison of the DETECTR^®^ assay to SARS-CoV-2 Whole-Genome Sequencing. **a**, MA plots, transformed onto M (log ratio) and A (mean average) scales, show CasDx1 SNP detection replicates (n = 807) for each SARS-CoV-2 mutation across 91 clinical samples. WT is denoted by blue dots, MUT is denoted by green dots, NoCall is denoted by orange dots and NTC is denoted by grey dots. **b**, Alignment of final mutation calls comparing the DETECTR^®^ and SARS-CoV-2 WGS assay results across 91 clinical samples after discordant samples (indicated by red asterisk) were resolved. **c**, Final lineage classification on each clinical sample by the DETECTR^®^ assay compared to the SARS-CoV-2 lineage determined by viral WGS. **d**, Final Positive Predictive Agreement (PPA), Negative Predictive Agreement (NPA) and concordance values for each WT and MUT SNP from the evaluation of the DETECTR^®^ assay against the SARS-CoV-2 WGS comparator assay after discordant samples were resolved.

We then unblinded the viral WGS results to evaluate the accuracy of the DETECTR^®^ assay for SNP calls and lineage classification. There were 14 discordant SNP calls out of 272 (94.9% SNP concordance) distributed among 11 clinical samples out of 91 (Extended Data Fig. 8a-c). Among the 11 discordant samples, one sample (COVID-31) was designated a ‘no call’ at position 452 by viral WGS and thus lacked a comparator, two samples were designated a ‘no call’ due to flat WT and MUT curves (COVID-41 and COVID-73), four samples had similar WT and MUT curve amplitudes, suggesting a mixed population (COVID-03, COVID-56, COVID-61 and COVID-81) (Extended Data Fig. 8a), and four samples had SNP assignments discordant with those from viral WGS (COVID-12, COVID-13, COVID-20 and COVID-63) (Extended Data Fig. 8a).

Given that the comparison data had been collected over an extended time period, we surmised that sample stability issues arising from aliquoting and multiple freeze-thaw cycles may have accounted for the observed discrepancies. To further investigate this possibility, the 11 discordant clinical samples were re-extracted from the original respiratory swab matrix and re-analyzed by running both viral WGS and the DETECTR^®^ assay in parallel. Re-testing of the samples resulted in nearly complete agreement between the two methods, except for two SNPs that were identified as E484Q in two samples by WGS but were incorrectly called E484 (WT) by the DETECTR^®^ assay (Fig. 3b-c **and** Extended Data Fig. 8d). Thus, based on discrepancy testing, the positive predictive agreement (PPA) between the DETECTR^®^ assay and viral WGS at all three WT and MUT SNP positions was 100% (272 of 272, p<2.2e-16 by Fisher’s Exact Test) (Fig. 3d). The corresponding negative predictive agreement (NPA) was 91.4% as the E484Q mutation for two SNPs was incorrectly classified as WT. Nevertheless, the final viral lineage classification for the 91 samples after discrepancy testing showed 100% agreement with viral WGS (Fig. 3d **and Supplementary Table 1**).

In November 2021, a new SARS-CoV-2 variant was identified and almost immediately designated a variant of concern, called Omicron^39^. The Omicron variant carries an exceptionally high number of mutations (>30) within the S-gene and has been shown to have enhanced transmissibility and immune evasion^40, 41^. The record number of COVID-19 cases globally from Omicron and loss of activity by certain therapeutic antibodies underscores the need for rapid and targeted identification of SARS-CoV-2 variants. Although the TaqPath PCR assay with S-gene Target Failure (SGTF) has functioned as a screen that can be reflexed to sequencing to identify the Omicron variant^42^, the SGTF assay alone cannot differentiate between Omicron BA.1 and Alpha^43, 44^ and cannot identify emerging variants that lack the SGTF, such as the Omicron BA.2 sublineage^43^. We therefore quickly reconfigured our COVID-19 variant DETECTR^®^ assay for the identification of Omicron by targeting the E484A mutation, which alone differentiates Omicron from all other current VBM/VOI/VOC. Given that E484-related mutations are present in multiple circulating variants and have a strong effect on reducing antibody neutralization^10, 45, 46^, we further updated our panel of CasDx1 gRNAs to detect all relevant mutations (E, K, Q and A) at amino acid position 484 (Fig. 4a-b).

**Fig. 4.**
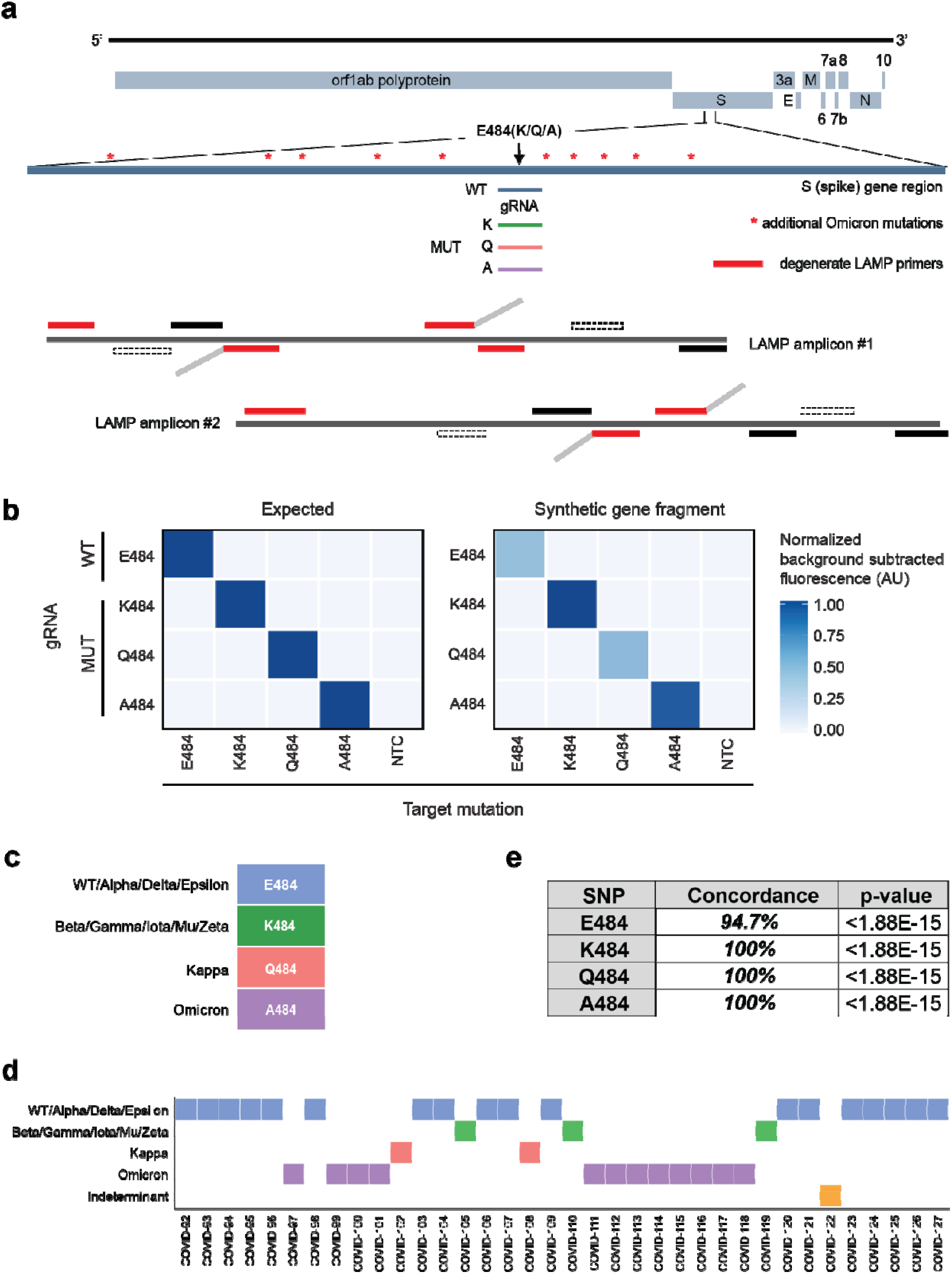
Specific detection of 484 mutations enables rapid Omicron identification. **a**, Schematic of Omicron mutations within the S-gene LAMP amplicon and relative position of 484-specific gRNAs and degenerate LAMP primers. **b**, Heat map comparison of end-point fluorescence (t = 30 min) showing specific detection of 484-specific mutations (E, K, Q, A) on PCR-amplified synthetic gene fragments (n = 3). **c**, SARS-CoV-2 lineage classification table based on 484 mutations. **d,** Alignment of final 484 mutation calls comparing the DETECTR^®^ and SARS-CoV-2 WGS assay results across 36 clinical samples. **e**, Overall SNP concordance values for the 484 SNP from the evaluation of the DETECTR^®^ assay against the SARS-CoV-2 WGS comparator assay.

Given the highly mutated Omicron S-gene, we suspected that our original LAMP primer set would not have sufficient sensitivity to amplify the targeted spike RGD region and thus we incorporated degenerate nucleotides within the LAMP primers to enable amplification of the Omicron S-gene (Fig. 4a, Extended Data Fig. 9**, and Supplementary Table 3**). Within weeks of the first Omicron case identified in the U.S.^47^, we procured and tested an additional set of 48 clinical samples. These samples were blinded and processed with the updated DETECTR^®^ assay workflow, which included sample extraction, followed by amplification with the degenerate LAMP primers and detection with each of the 484-specific CasDx1 gRNAs. Once processed, a result with mutations K484, Q484 or A484 was called Beta/Eta/Gamma/Iota/Mu/Zeta, Kappa, or Omicron, respectively (Fig. 4c-e **and** Extended Data Fig. 10a). If the result was associated with E484 (WT), we ran the assay using WT and MUT gRNAs at positions 452 and 501 to call the final SARS-CoV-2 lineage (Fig. 4c-e **and** Extended Data Fig. 10a). Using this workflow, we detected 36 out of 48 total clinical samples: 18/48 resulted as E484 (WT) and were subsequently tested with 452 and 501 gRNAs (3/18 called WT, 6/18 called Alpha and 9/18 called Delta), 4/48 resulted as K484 (called Beta/Eta/Gamma/Iota/Mu/Zeta), 2/48 resulted as Q484 (called Kappa), and 12/48 resulted as A484 (called Omicron) (Fig. 4e **and** Extended Data Fig. 10b-c). The remaining 12/48 clinical samples neither amplified nor showed any DETECTR^®^ signal and were thus called “Not Detected” (**Supplementary Table 2**).

Unblinding of samples COVID-92 through COVID-127 revealed five discordant samples: COVID-103, COVID-108, COVID-109, COVID-112 and COVID-122 (Extended Data Fig. 10d). All five discordant samples were re-extracted from the original patient sample and re-processed with WGS and COVID Variant DETECTR^®^. After repeat testing, three samples (COVID-103, COVID-108, COVID-109) showed 100% concordance between WGS and DETECTR^®^, with both methods resulting in “No Call” at position 452. Notably, these samples were also part of the original set of 91 samples (COVID-20, COVID-63, COVID-73) that were previously concordant at position 452, suggesting a decrease in sample integrity likely resulting from multiple freeze/thaw cycles incurred during several re-extractions. Sample COVID-112 was called an Omicron by DETECTR^®^ based on its A484 SNP call, which was confirmed by WGS. Finally, sample COVID-122 could not be amplified by RT-LAMP, also suggesting a loss in sample integrity. Following this discrepancy analysis, we demonstrated an overall SNP concordance of 94.7%, and 100% NPA for this set of 48 samples (Fig. 4e).

## Discussion

In this study, we developed a CRISPR-based DETECTR^®^ assay for the detection of SARS-CoV-2 variants. We evaluated three CRISPR-Cas12 enzymes, two commercially available (LbCas12a from NEB and AsCas12a from IDT) and one proprietary (CasDx1 from Mammoth Biosciences). Based on a head-to-head comparison of these enzymes, we observed clear differences in performance, with CasDx1 demonstrating the highest fidelity as the only enzyme able to reliably detect all targeted SNPs. A data analysis pipeline, developed to differentiate between WT and MUT signals with the DETECTR^®^ assay, yielded an overall SNP concordance of 97.9% (373/381 total SNP calls) and 99.3% (138/139) agreement with lineage classification compared to viral WGS. These findings show robust agreement between the DETECTR^®^ assay and viral WGS for identification of SNP mutations and variant categorization. Thus, the DETECTR^®^ assay provides a faster and simpler alternative to sequencing-based methods for COVID-19 variant diagnostics and surveillance.

Our results show that the choice of Cas enzyme is important to maximize the accuracy of CRISPR-based diagnostic assays and may need to be tailored to the site that is being targeted. As currently configured with the L452R, E484K/Q/A and N501Y SNP targets, the COVID-19 variant DETECTR^®^ assay is currently capable of distinguishing the Alpha, Delta, Kappa and Omicron variants, but cannot resolve the remaining VBMs or VOIs. However, given the rapid emergence and shifts in the distribution of variants over time^13^ it is likely that tracking of key mutations, many of which are suspected to arise from convergent evolution^48^, rather than tracking of variants, will be more important for surveillance as the pandemic continues. Here we also developed a data analysis pipeline for CRISPR-based SNP calling that can readily incorporate additional targets and offers a blueprint for automated interpretation of fluorescent signal patterns.

Although CRISPR-based diagnostic assays have been previously demonstrated for the detection of SARS-CoV-2 variants, these studies have limitations regarding coverage of circulating lineages, the extent of clinical sample evaluation, and/or assay complexity. For example, the miSHERLOCK variant assay uses LbCas12a (NEB) with RPA pre-amplification to detect N501Y, E484K and Y144Del covering eight lineages (WA-1, Alpha, Beta, Gamma, Eta, Iota, Mu and Zeta) and was tested only on contrived samples (RNA spiked into human saliva)^21^. The SHINEv2 assay uses LwaCas13a with RPA pre-amplification to detect 69/70Del, K417N/T, L452R and 156/157Del + R158G covering eight lineages (WA-1, Alpha, Beta, Gamma, Delta, Epsilon, Kappa and Mu) and was tested with only the 69/70Del gRNAs on 20 Alpha-positive NP clinical samples^49^. Finally, the mCARMEN variant identification panel (VIP) uses 26 crRNA pairs with either the LwaCas13a or LbaCas13a and PCR pre-amplification to identify all current circulating lineages including Omicron; however, the VIP requires the Fluidigm Biomark HD system or similar, more complex instrumentation for streamlined execution^50^. In comparison, the DETECTR^®^ assay presented here uses CasDx1 with LAMP pre-amplification to detect N501Y, E484K/Q/A and L452R covering all current circulating lineages including Omicron and tested on 139 clinical samples representing eight lineages (WA-1, Alpha, Gamma, Delta, Epsilon, Iota, Mu, Omicron). Furthermore, we demonstrate here that specific Omicron identification can be accomplished using only the E484 WT and A484 MUT guides.

Some limitations of our study are as follows. First, as previously mentioned, the DETECTR^®^ assay currently detects only the L452R, E484K/Q/A and N501Y mutations, which may not provide enough resolution to identify future lineages. Second, we observed variable performance of the assay in SNP discrimination, with more potential overlap in the calls between WT and MUT for the 452 position than for the other two sites. These two limitations could potentially be addressed by the incorporation of additional gRNAs to the assay to provide specific and redundant coverage and to improve identification of specific lineages. Third, due to a multiplexed and degenerate S-gene LAMP primer design, the limit of detection of the DETECTR^®^ assay is higher than our previously published SARS-CoV-2 DETECTR^®^ assay^20^, and thus only positive clinical samples with a Ct < 30 (near the limit of detection for viral whole-genome sequencing) were tested in our study. Incorporation of an additional N-gene target to the assay may be necessary if simultaneous detection and SNP/variant identification is desired. Finally, the current study focuses on the development and validation of a variant DETECTR^®^ assay using conventional laboratory equipment. Future work will involve implementation onto automated, portable systems for use in point of care settings.

In the near term, we suggest the use of the DETECTR^®^ assay as an initial screen for circulating variants and/or a distinct pattern from a rare or novel variant by interrogating the key 452, 484, and 501 positions that could be reflexed to viral WGS. As the sequencing capacity for most clinical and public health laboratories is limited, the DETECTR^®^ assay would thus enable rapid identification of variants circulating in the community to support outbreak investigation and public health containment efforts. Identification of specific mutations associated with neutralizing antibody evasion^12, 51^ could inform patient care with regards to the use of monoclonal antibodies that remain effective in treating the infection^15^. As the virus continues to mutate and evolve, the DETECTR^®^ assay can be readily reconfigured by validating new gRNAs and pre-amplification LAMP primers and gRNAs that target emerging mutations with clinical and epidemiological significance. Over the longer term, a validated CRISPR assay that combines SARS-CoV-2 detection with variant identification offers a faster and simpler alternative to sequencing and would be useful as a tool for simultaneous COVID-19 diagnosis in individual patients and surveillance for infection control and public health purposes.

## Materials and Methods

### Synthetic Gene Fragments

Wild-type (WT) and mutant (MUT) synthetic gene fragments (Twist) were PCR amplified using NEB 2x Phusion Master Mix following the manufacturer’s protocol. The amplified product was cleaned using AMPure XP beads following manufacturers protocol at a 0.7x concentration. The product was eluted in nuclease-free water and normalized to 10 nM. All nucleic acids used in this study are summarized in Supplementary Table 3.

### Clinical sample acquisition and extraction

De-identified residual SARS-CoV-2 RT-PCR positive nasopharyngeal and/or oropharyngeal (NP/OP) swab samples in universal transport media (UTM) or viral transport media (VTM) were obtained from the UCSF Clinical Microbiology Laboratory. All samples were stored in a biorepository according to protocols approved by the UCSF Institutional Review Board (protocol number 10-01116, 11-05519) until processed.

All NP/OP swab samples obtained from the UCSF Clinical Microbiology Laboratory were pretreated with DNA/RNA Shield (Zymo Research, # R1100-250) at a 1:1 ratio. The Mag-Bind Viral DNA/RNA 96 kit (Omega Bio-Tek, # M6246-03) on the KingFisher Flex (Thermo Fisher Scientific, # 5400630) was used for viral RNA extraction using an input volume of 200 μl of diluted NP/OP swab sample and an elution volume of 100 μl. The Taqpath™ COVID-19 RT-PCR kit (Thermo Fisher Scientific) was used to determine the N gene cycle threshold values.

### Heat-inactivated culture acquisition and extraction

Heat-inactivated cultures of SARS-CoV-2 Variants Being Monitored (VBM), Variants of Concern (VOC) or Variants of Interest (VOI) were provided by the California Department of Public Health (CDPH).

RNA from heat-inactivated SARS-CoV-2 VBM/VOC/VOI isolates were extracted using the EZ1 Virus Mini Kit v2.0 (Qiagen, # 955134) on the EZ1 Advanced XL (Qiagen, # 9001875) according to the manufacturer’s instructions. For each culture, six replicate LAMP reactions were pooled into a single sample. DETECTR^®^ was performed on a 1:10 dilution of the 10,000 cp/rxn LAMP amplification products.

### COVID-19 variant DETECTR^®^ assay

Two LAMP primer sets, each containing 6 primers, were designed to target the L452R, E484K and N501Y mutations in the SARS-CoV-2 Spike (S) protein (Supplemental Table). Sets of LAMP primers were designed from a 350 bp target sequence spanning the 3 mutations using Primer Explorer V5 (https://primerexplorer.jp/e/). Candidate primers were manually evaluated for inclusion using the OligoCalc online oligonucleotide properties calculator^52^ while ensuring that there was no overlap with either primers from the other set or guide RNA target regions that included the L452R, E484K, and N501Y mutations.

Multiplexed RT-LAMP was performed using a final reaction volume of 50 μl, which consisted of 8 μl RNA template, 5 μl of L452R primer set (Eurofins Genomics), 5 μl of E484K/N501Y primer set, 17 μl of nuclease-free water, 1 μl of SYTO-9 dye (ThermoFisher Scientific), and 14 μl of LAMP mastermix. Each of the primer sets consisted of 1.6 μM each of inner primers FIP and BIP, 0.2 μM each of outer primers F3 and B3, and 0.8 μM each of loop primers LF and LB). The LAMP mastermix contained 6 mM of MgSO4, isothermal amplification buffer at 1X final concentration, 1.5 mM of dNTP mix (NEB), 8 units of Bst 2.0 WarmStart DNA Polymerase (NEB), and 0.5 ul of WarmStart RTx Reverse Transcriptase (NEB). Plates were incubated at 65°C for 40 minutes in a real-time Quantstudio^™^ 5 PCR instrument. Fluorescent signals were collected every 60 seconds

Degenerate multiplexed RT-LAMP was performed using a final reaction volume of 65 μl, which consisted of 9.6 μl RNA template, 10 μl of L452R degenerate primer set (Eurofins Genomics), 10 μl of E484K/N501Y degenerate primer set, 14.1 μl of nuclease-free water, 1.3 μl of SYTO-9 dye (ThermoFisher Scientific), and 20 μl of LAMP mastermix.The primer set and mastermix assembly, the incubation and data collection were described above.

40nM CasDx1 (Mammoth Biosciences), LbCas12a (EnGen^®^ Lba Cas12a, NEB) or AsCas12a (Alt-R^®^ A.s. Cas12a, IDT) protein targeting the WT or MUT SNP at L452(R), E484(K) or N501(Y) was incubated with 40nM gRNA in 1X buffer (MBuffer3 for CasDx1, NEBuffer r2.1 for LbCas12a and AsCas12a) for 30 min at 37°C. Dx1 gRNAs were used with both CasDx1 and LbCas12a, whereas AsCas12a gRNAs were used with AsCas12a (Supplementary Table 3). 100nM ssDNA reporter (/5Alex594N/TTATTATT/3IAbRQSp/, IDT) was added to the RNA-protein complex. 18μL of this DETECTR® master mix was combined with 2 μL target amplicon. The DETECTR® assays were monitored for 30 min at 37oC in a plate reader (Tecan).

### Digital PCR

Samples were evaluated at 3 dilutions (1:100; 1:1,000; and 1:10,000) using the ApexBio Covid-19 Multiplex Digital PCR Detection Kit (Stilla Technologies) according to the manufacturer’s protocol. The controls (positive and negative provided by UCSF, the Kit Controls, and an internal control) were run with the samples in duplicate. The dilutions were used to determine the most accurate concentration which was determined from the N gene concentration.

### Sequencing methods

Complementary DNA (cDNA) synthesis from RNA via reverse transcription and tiling multiplexed amplicon PCR were performed using SARS-CoV-2 primers version 3 according to the Artic protocol^53, 54^. Libraries were constructed by ligating adapters to the amplicon products using NEBNext Ultra II DNA Library Prep Kit for Illumina (New England Biolabs, # E7645L), barcoding using NEBNext Multiplex Oligos for Illumina (New England Biolabs, # E6440L), and purification with AMPure XP (Beckman-Coulter, # 63880). Final pooled libraries were sequenced on either Illumina Miseq or NextSeq 550 as 2×150 single-end reads (300 cycles).

SARS-CoV-2 viral genome assembly and variant analyses were performed using an in-house bioinformatics pipeline. Briefly, sequencing reads generated by Illumina sequencers (MiSeq or NextSeq 550) were demultiplexed and converted to FASTQ files using bcl2fastq (v2.20.0.422). Raw FASTQ files were first screened for SARS-CoV-2 sequences using BLASTn (BLAST+ package 2.9.0) alignment against the Wuhan-Hu-1 SARS-CoV-2 viral reference genome (NC_045512). Reads containing adapters, the ARTIC primer sequences, and low-quality reads were filtered using BBDuk (version 38.87) and then mapped to the NC_045512 reference genome using BBMap (version 38.87). Variants were called with CallVariants and iVar (version 1.3.1) and a depth cutoff of 5 was used to generate the final assembly. Pangolin software (version 3.1.17)^55, 56^ was used to identify the lineage. Using a custom in-house script, consensus FASTA files generated by the genome assembly pipeline were scanned to confirm L452R, E484K, and N501Y mutations.

### Discordant sample retesting

The discordant samples (n=16) were re-extracted as described above for the NP/OP swab samples and evaluated by viral WGS as described above. The extracted nucleic acids were then thawed (incurring an additional freeze/thaw as needed) and amplified using the LAMP protocol described above and evaluated using the DETECTR**^®^** assay as described above.

### DETECTR**^®^** data analysis pipeline

#### Quality Control Metric for the LAMP Reaction

Prior to processing DETECTR^®^ data from the clinical samples, we collected data indicating the success or failure of the samples to amplify in the LAMP reaction. The absolute truth was based on visual inspection of LAMP curves This absolute truth was used to develop thresholds for the LAMP reactions. The positive and negative controls from the LAMP reactions were used to derive the thresholds to qualify the samples. Two sets of thresholds were used: time threshold and fluorescence rate threshold. The positive LAMP controls were assumed to represent an ideal sample and displayed a classic sigmoidal rise of fluorescence over time and the NTC represented the background fluorescence. It was hypothesized that a sample will ideally have positive control like fluorescence kinetics. However, due to the presence of high background in some samples, a mean value between controls for each plate was chosen as threshold. After this, the fluorescence values at a time threshold of 18 minutes were collected. The time point is of importance here to rule out those samples that would amplify closer to the endpoint, signifying the LAMP intermediates to be the majority contributors of the rise in the signal and not the actual sample itself. A score was assigned for each sample which was calculated as a ratio of rate of fluorescence rate threshold to the rate of fluorescence value at 18 minutes for each sample. The hypothesis is that if this ratio of rate of fluorescence between controls and samples is less than 1, then samples have failed to reach the minimum fluorescence required to be called out as amplified and if the ratio is greater than or equal to 1, then samples have amplified sufficiently. To identify the exact score value for a qualitative QC metric, an ROC analysis was done on scores and the absolute truth (Extended Data Fig. 6).

#### Data Analysis for CRISPR-based SNP calling

Each well had a guide specific to the mutant or the wild-type SNP. The comparison is important to assign a genotypic call to the sample. The DETECTR^®^ reactions across the plate are not comparable to each other. For this purpose, the endpoint fluorescence intensities are normalized in each well to its own minimum intensity. This term is called fluorescence yield. The fluorescence yield was compared across wells in a plate under the assumption that each well would have similar minimum fluorescence starting point. Irrespective of the highest levels of the fluorescence intensities observed across samples, the yield for a given target must ideally remain the same assuming that similar concentrations of samples/target are being compared. This aided in normalizing the signal and comparing replicates across the wells in the same plate.

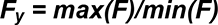

The wild-type and mutant target guides on NTC must ideally not show any change in intensity over time. The fluorescence yield for NTC must remain constant across replicates, plates and close to 1.

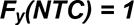

On the contrary, if a sample has a fluorescence yield of 1, then it qualifies for a No Call.

#### General rules for variant calling

1. NTC was assigned NTC
2. If the Contrast of the sample for a SNP was between minimum and maximum contrast for the plate, then the sample is assigned a NoCall.
3. If the Size of the sample is lower than the Size of the NTC on the plate, then the sample is assigned a NoCall.

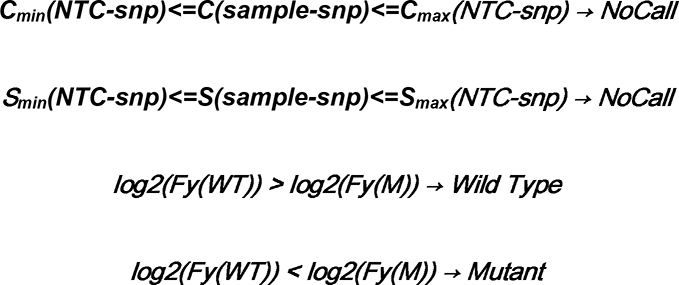

In cases, where more than one mutant at a particular position was being analyzed for, then the signals of each mutant was compared with the wild-type signal. If there existed a mutant, then among (n) comparisons for n mutants and one wild type, one of the comparisons would yield mutant call.

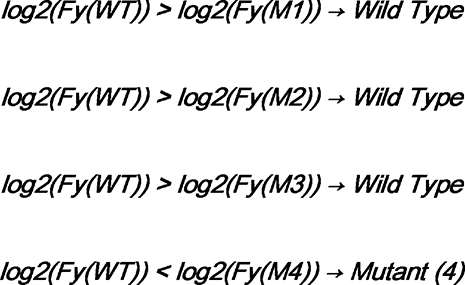

If there was a tie in the above logic between mutant and wild-type, then a tie breaker comparison would yield a final result.

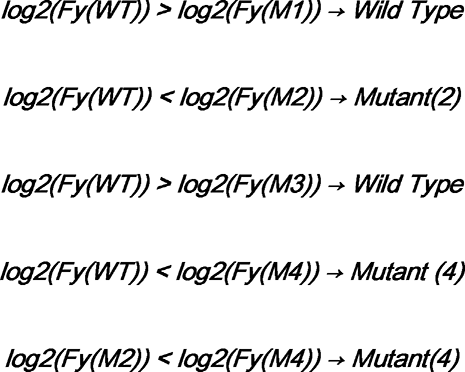

#### SNP Calls

We used the following procedure to evaluate the concordance between sequencing and DETECTR^®^ technologies for genotypic classification of the clinical cohort dataset.

First, we considered all samples and SNPs for which both sequencing and DETECTR^®^ data was present in the distributed files by matching the SNP IDs and sample names. This included cleaning and curing the dataset which had failed LAMP reactions and identifying WT and MUT based on the spacer fluorescent. This yielded a preliminary data set containing 279 calls across three SNPs against 93 samples. After eliminating samples that had failed to amplify in the LAMP reaction but were assigned a genotype, the resulting final analysis data consisted of 272 calls (WT, MUT and NoCall) spread across three SNPs and 91 samples. For each of the three SNPs in the analysis data set, we identified and recorded both sequencing and DETECTR^®^ genotypes (including NoCalls and LAMP Fails) for each of the 93 patients. The 91 patients include the individuals for whom actual sequencing data was available

### Statistical analysis

#### SNP Calls

For each SNP in the analysis, we computed a variety of statistics evaluating the concordance between genotype calls on the two different technologies. The concordant and discordant genotypes were visualized through contingency tables. For each SNP, there are three possible genotypes (WT, MUT and No Call). The concordance rates were calculated without the samples that failed the LAMP reaction (Fig. 3B **and** Supplementary Table 1). The 2×2 cross tables classify all three SNPs across all the samples between sequencing and DETECTR^®^ technologies (Fig. 3B **and** Supplementary Table 1). The data transformation and statistical analysis was done in R^57^.

## Data Availability

All data needed to evaluate the conclusions in the paper are present in the paper and/or the Supplementary Materials. The CasDx1 protein can be provided by Mammoth Biosciences to the extent feasible, pending scientific review and a completed material transfer agreement. Requests for the CasDx1 protein should be submitted to Janice Chen at.

## Acknowledgments

We thank the UCSF Center for Advanced Technology core facility (Delsy Martinez and Tyler Miyasaki) for their efforts in high-throughput sequencing of viral cDNA libraries using the Illumina NovaSeq 6000 instrument, and Mary Kate Morris from the California Department of Public Health for providing the heat-inactivated viral cultures. We also thank Lucas Harrington and Teresa Peterson for their critical review of this manuscript.

This work has been funded by the Innovative Genomics Institute (IGI) at UC Berkeley and UC San Francisco (C.Y.C.), the Sandler Program for Breakthrough Biomedical Research (C.Y.C.), US Centers for Disease Control and Prevention contract 75D30121C10991 (C.Y.C.), and Mammoth Biosciences.

## Author contributions

C.L.F., J.S.C., and C.Y.C. conceived and designed the study. C.Y.C, and V.S. coordinated the SARS-CoV-2 whole-genome sequencing efforts and RT-LAMP primer design and testing. C.L.F., J.P.B., J.C., and J.S.C. designed guide RNAs for CRISPR-Cas12 testing. B.M., J.P.B., R.N.D., E.S., and C.G.H. tested guide RNAs and ran DETECTR^®^ experiments. V.S., N.B., B.W., A.S.-G., K.R., J.S., S.M., and C.Y.C. collected samples. C.L.F., V.S., B.M., V.N., J.P.B., J.C., J.S.C., and C.Y.C. analyzed data. C.L.F., V.S., V.N., B.M., J.P.B., J.C., J.S.C., and C.Y.C. wrote the manuscript. All authors read the manuscript and agree to its contents.

## Supplementary Figures

**Extended Data Fig. 1.**
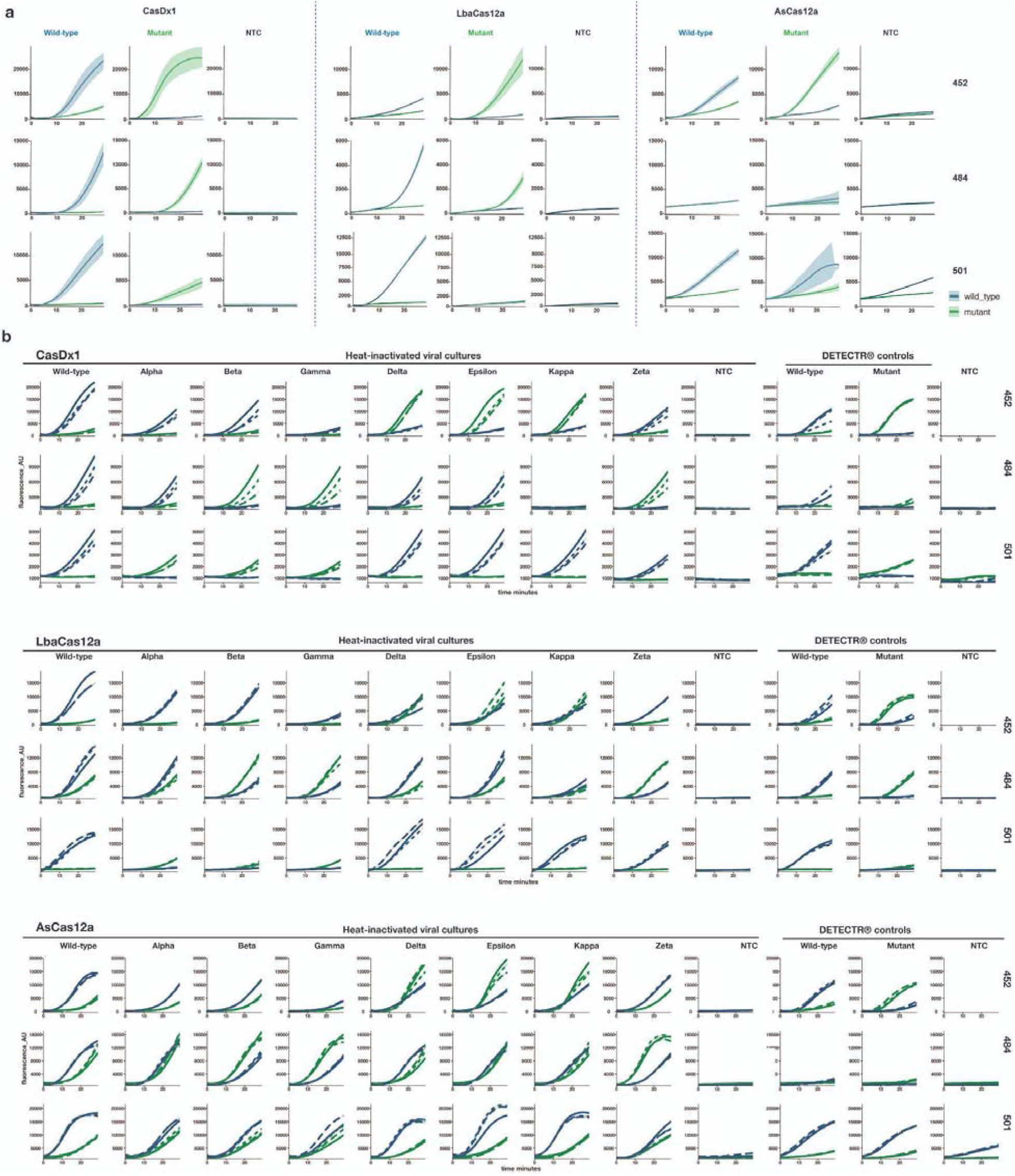
DETECTR^®^ curves from gene fragments and heat-inactivated viral cultures. **a**, Raw fluorescence curves from three Cas12 enzymes (CasDx1, LbCas12a, AsCas12a) complexed with WT and MUT SNP gRNAs run on PCR-amplified gene fragments representing WT and MUT SNP targets. **b**, Raw fluorescence curves from three Cas12 enzymes (CasDx1, LbCas12a, AsCas12a) on eight heat-inactivated viral culture samples from various SARS-CoV-2 lineages, a no target control (RT-LAMP) and CasDx1 detection controls (WT, MUT and NTC). CasDx1 replicates (n = 3), ±1.0SD.

**Extended Data Fig. 2.**
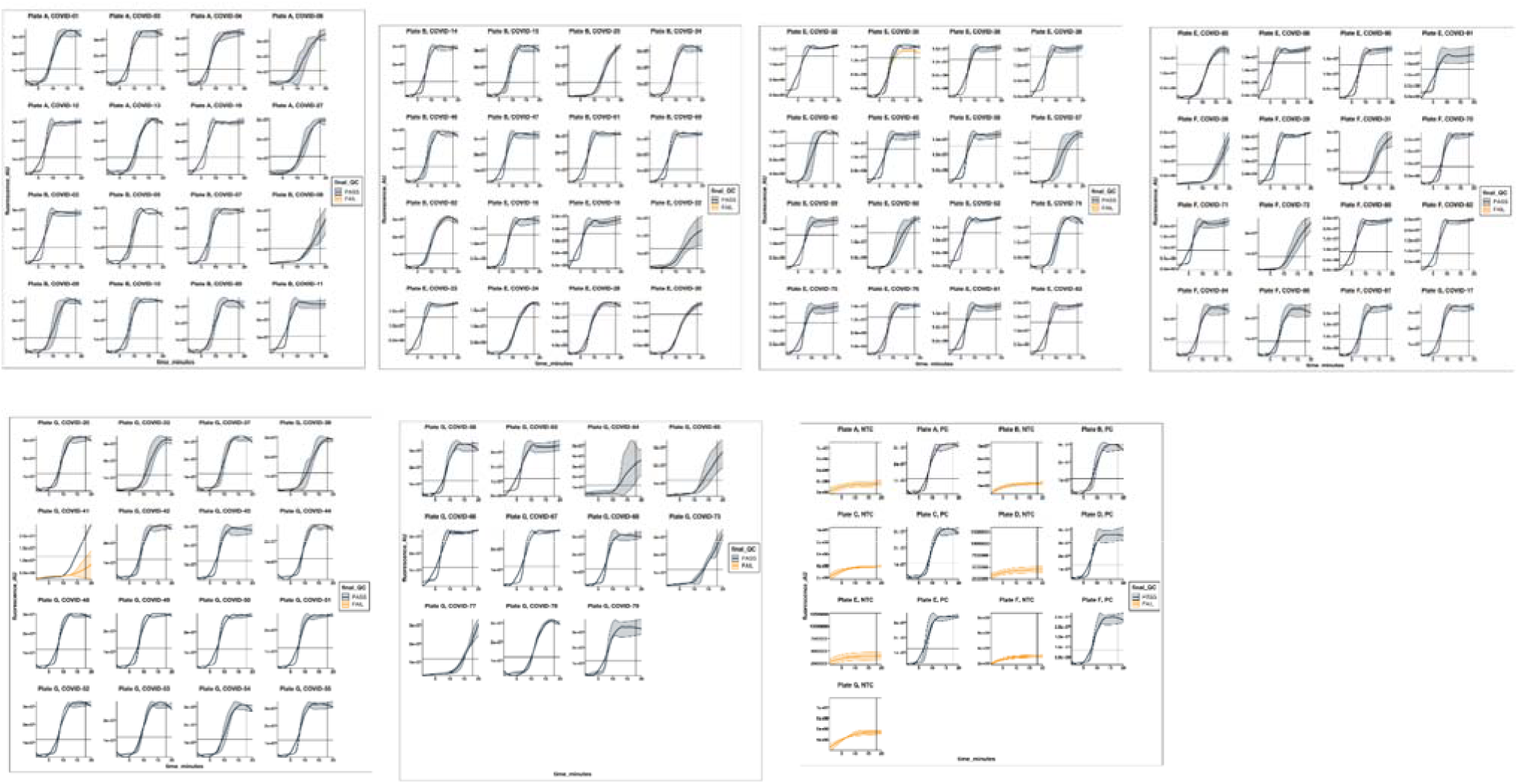
Raw fluorescence RT-LAMP curves for each clinical sample. The raw fluorescence RT-LAMP amplification curves for each of the clinical samples analyzed (n = 3 replicates). Each line is representative of the median ±1.0SD of the three RT-LAMP replicates for each sample. RT-LAMP replicates that passed QC are represented in navy blue and failed LAMP replicates are shown in orange. Only valid RT-LAMP replicates were used in subsequent data analysis.

**Extended Data Fig. 3.**
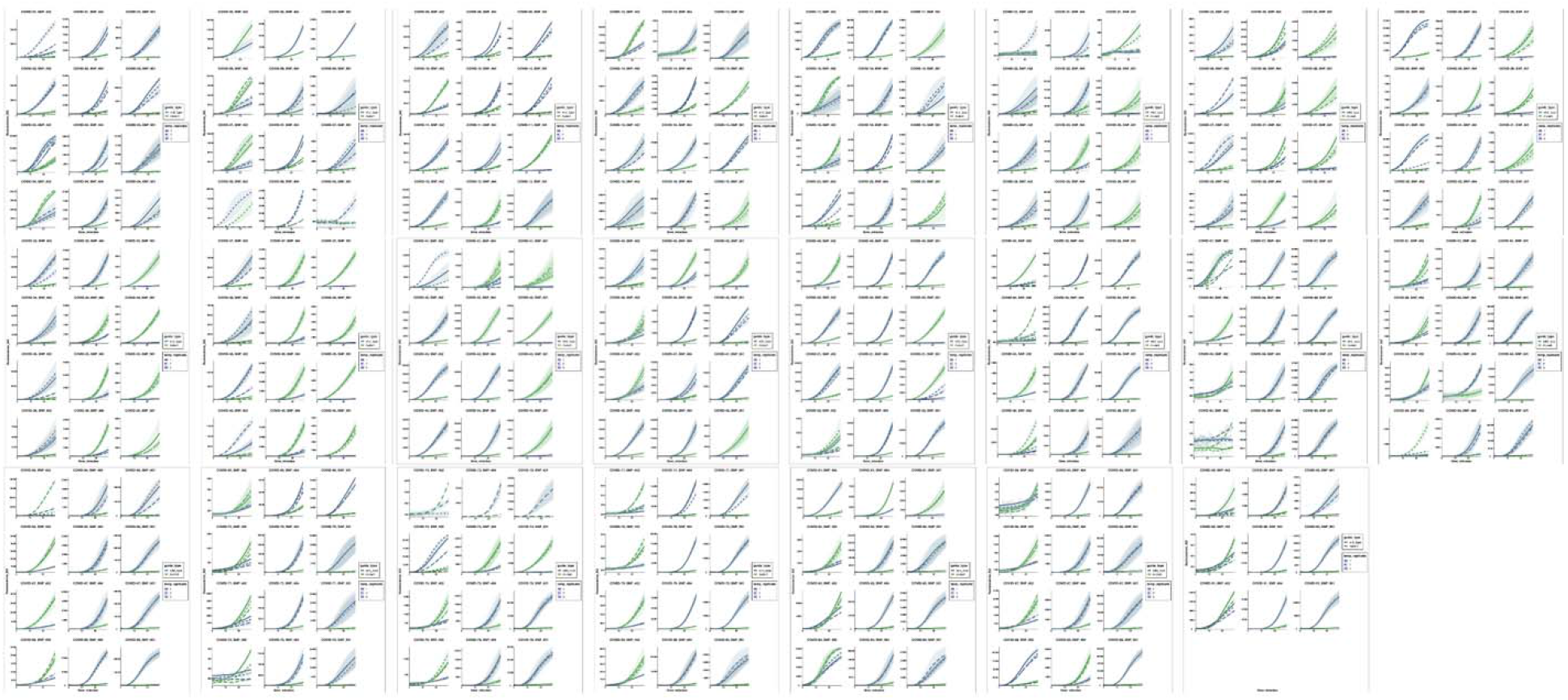
Raw fluorescence CasDx1 curves for each clinical sample amplified by RT-LAMP. Each clinical sample was amplified with RT-LAMP in triplicate, and the resulting amplicons were detected by CasDx1 in triplicate. The raw fluorescence curves show WT detection in blue and MUT detection in green. Each line is representative of the median ±1.0SD of the CasDx1 replicates (n = 3) for each WT and MUT guide for each of the RT-LAMP replicates (n = 3), represented by different patterned lines.

**Extended Data Fig. 4.**
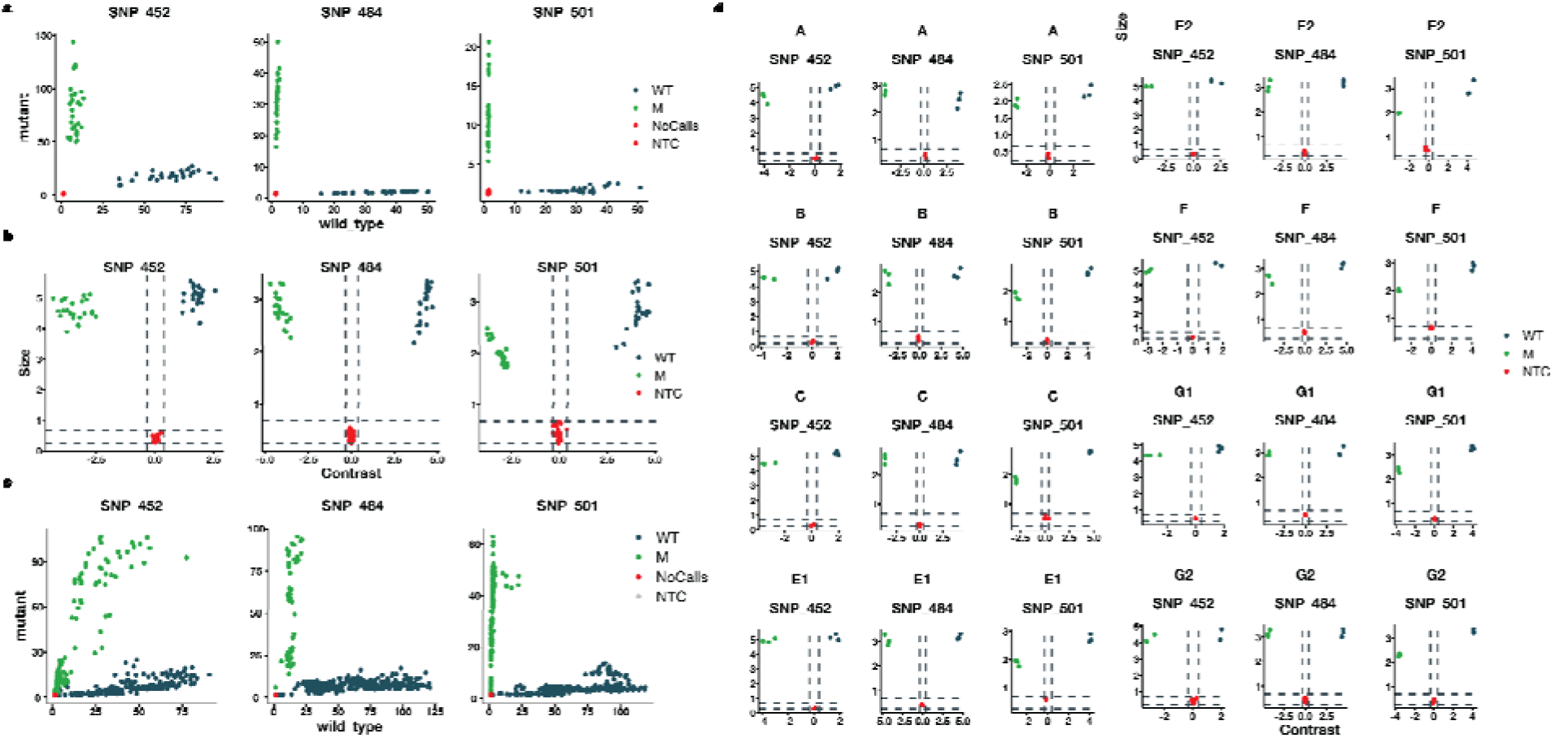
Evaluation of the DETECTR^®^ data analysis pipeline and making final calls. a, Allele discrimination plot visualizing the scaled signals from the COVID Variant DETECTR^®^ assay on gene fragments. The allele discrimination plots represent scatter plots of scaled WT and MUT fluorescence values plotted against each other. b, Contrast-Size plots of the COVID Variant DETECTR^®^ assay data on gene fragments to decrease ambiguity of the scaled signals, a ratio of the WT and MUT transformed values are plotted against the average of the WT and MUT transformed values on the MA plot. c, Allele discrimination plot visualizing the scaled signals from the COVID Variant DETECTR^®^ assay on clinical sample. d, MA plots of the COVID Variant DETECTR^®^ assay on the gene fragments (n = 30 WT; n = 30 MUT for each SNP) and no template controls (n = 33 WT; n = 33 MUT for each SNP) used to test the data analysis.

**Extended Data Fig. 5.**
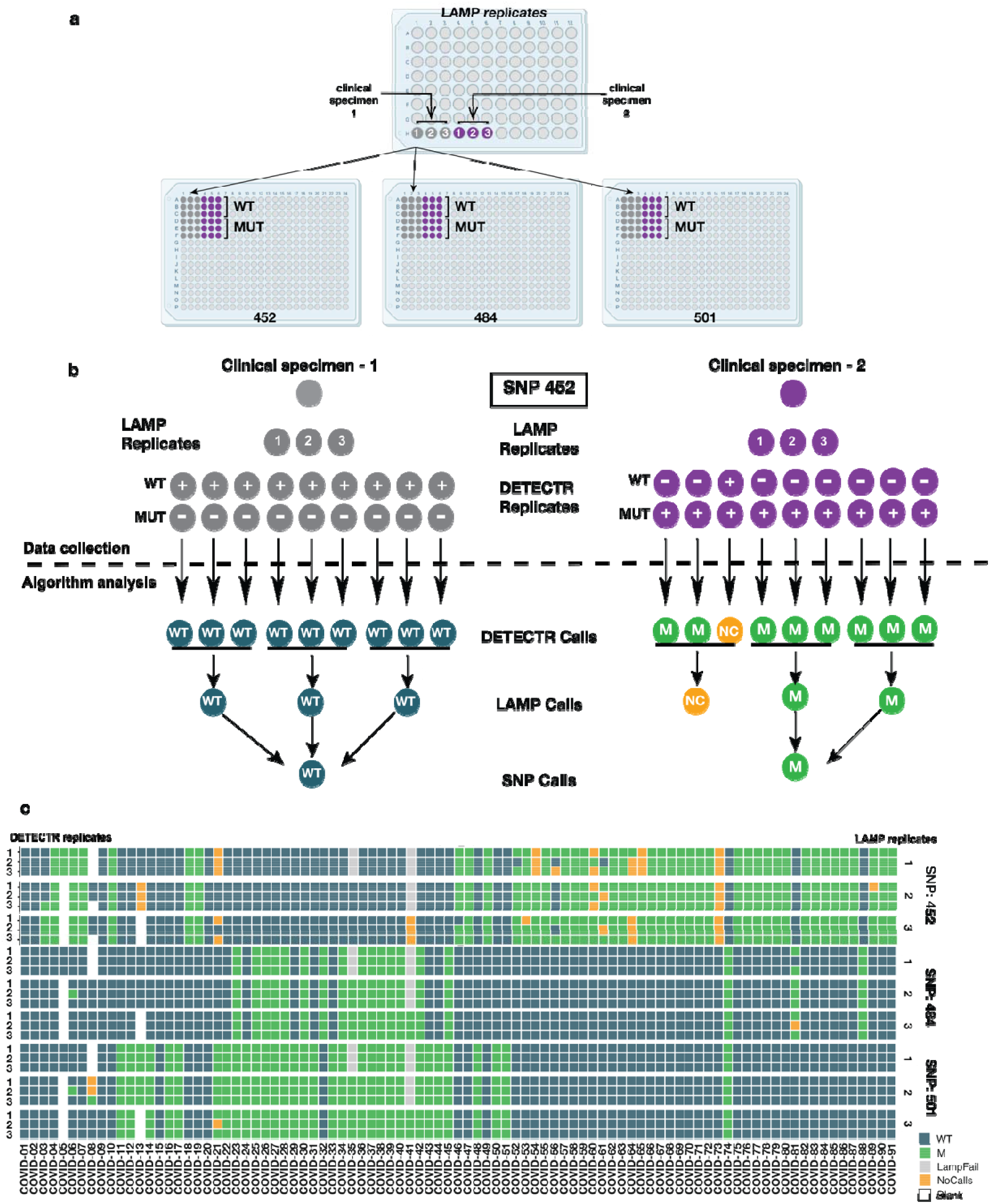
Highly specific detection by CasDx1 for each SNP on RT-LAMP replicates from clinical samples. **a,** The DETECTR^®^ assay workflow from LAMP amplification to SNP identification. **b**, Schematic showing the relationship between clinical samples, LAMP replicates and CasDx1 replicates that culminate in a final SNP call. **c**, Heat map showing CasDx1 signal (n = 3) per every LAMP replicate (n = 3) for each SNP on every clinical sample reflecting samples prior to discordance testing.

**Extended Data Fig. 6.**
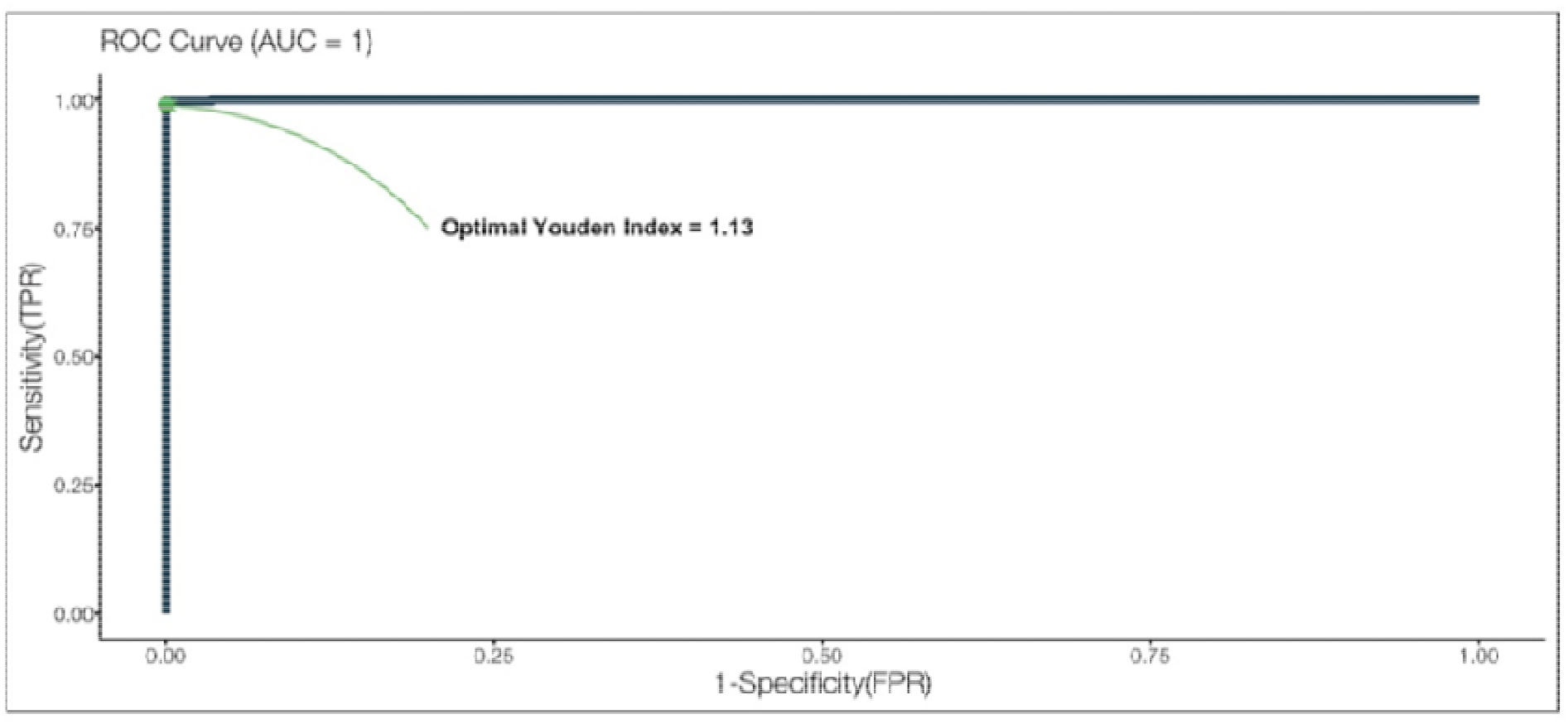
Determination of RT-LAMP threshold with a ROC curve. Thresholds for LAMP quality analysis were derived to determine which samples had amplified sufficiently. The exact score value for this qualitative QC metric was determined using a ROC analysis.

**Extended Data Fig. 7.**
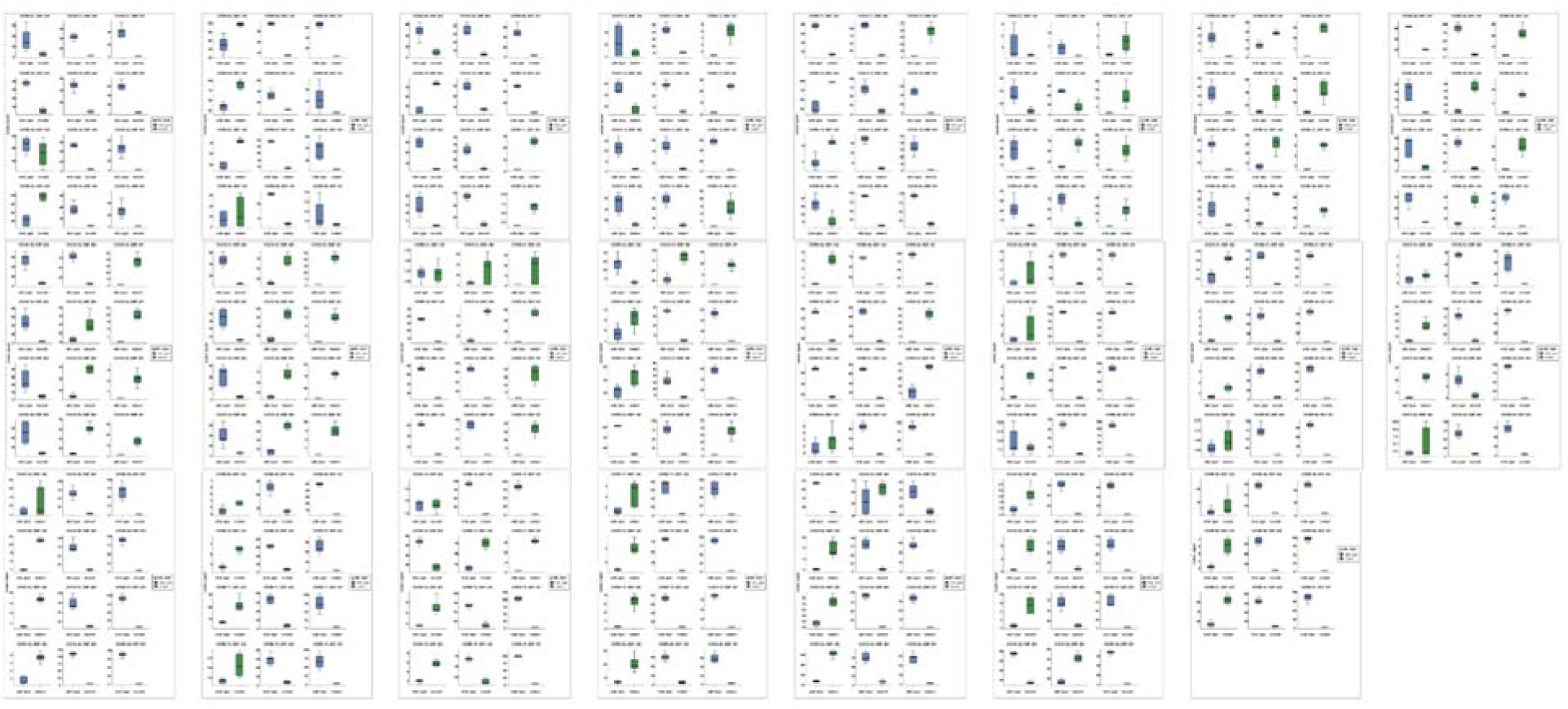
Visualization of SNP calls by the DETECTR^®^ data analysis pipeline. Box plots of all the clinical samples illustrate the spread of the scaled signals for each of the samples across the replicates in the experiment. SNP calls were made on each sample agreement with the median values depicted on the box plot of the sample, which also provided an analytical confirmation of the DETECTR^®^ results.

**Extended Data Fig. 8.**
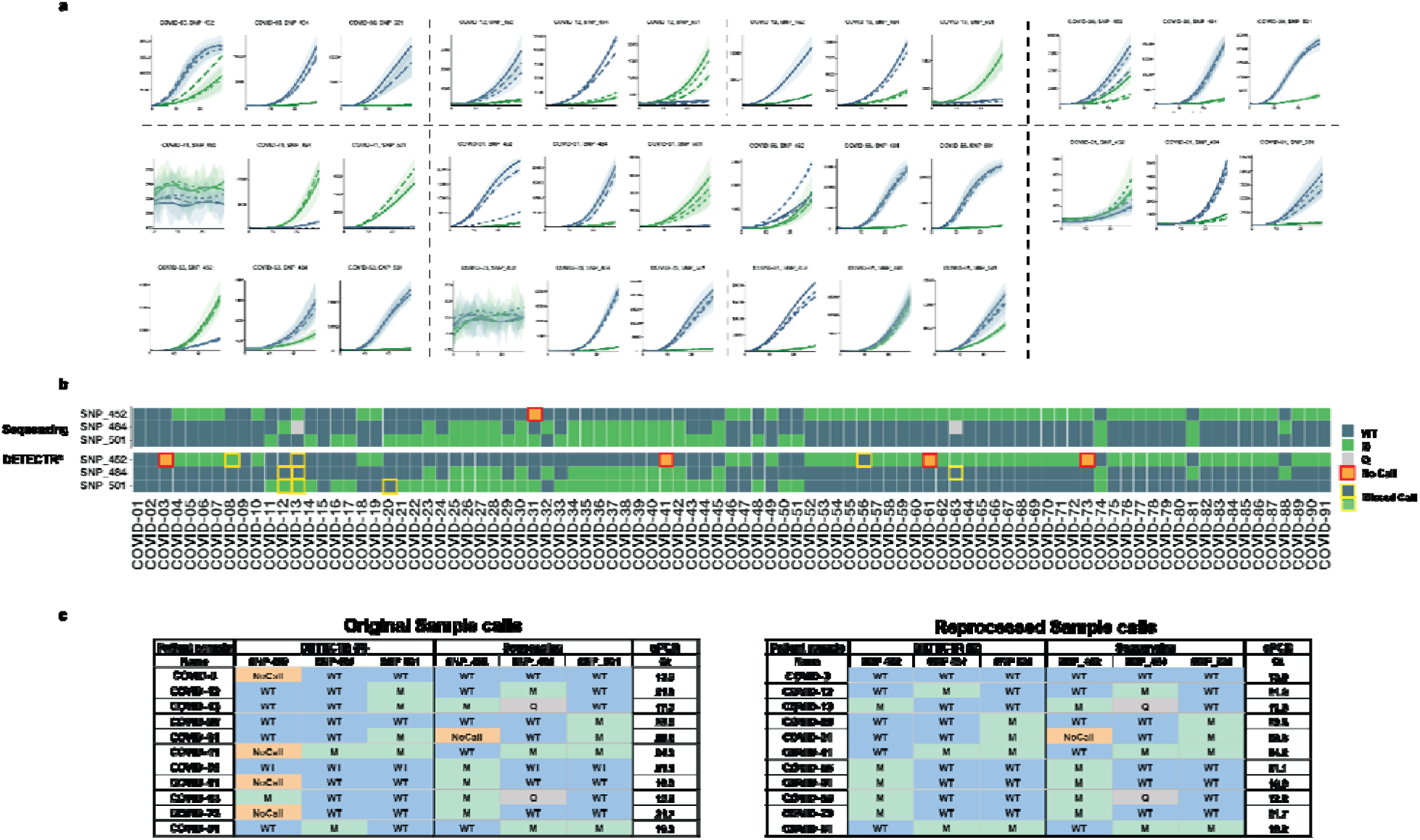
Clinical evaluation results with clinical samples of uncertain integrity. **a**, Raw fluorescence CasDx1 curves for the clinical samples with discordant DETECTR^®^ and WGS results. WT detection is represented by blue lines and MUT detection is represented by green lines. Each line is representative of the median ±1.0SD of the CasDx1 replicates (n = 3) for each guide for each of the LAMP replicates (n = 3), and each RT-LAMP replicate is represented by different patterned lines. **b**, Visualization of the COVID Variant DETECTR^®^ and SARS-CoV-2 WGS assays showing the alignment of final calls. Across all of the clinical samples in this cohort, 80 out of the 91 clinical sample COVID Variant DETECTR^®^ assay calls were consistent with the SARS-CoV-2 WGS calls. **c**, Summary of re-testing of discordant samples from the original clinical sample shows nearly all SNP discrepancies are resolved.

**Extended Data Fig. 9.**
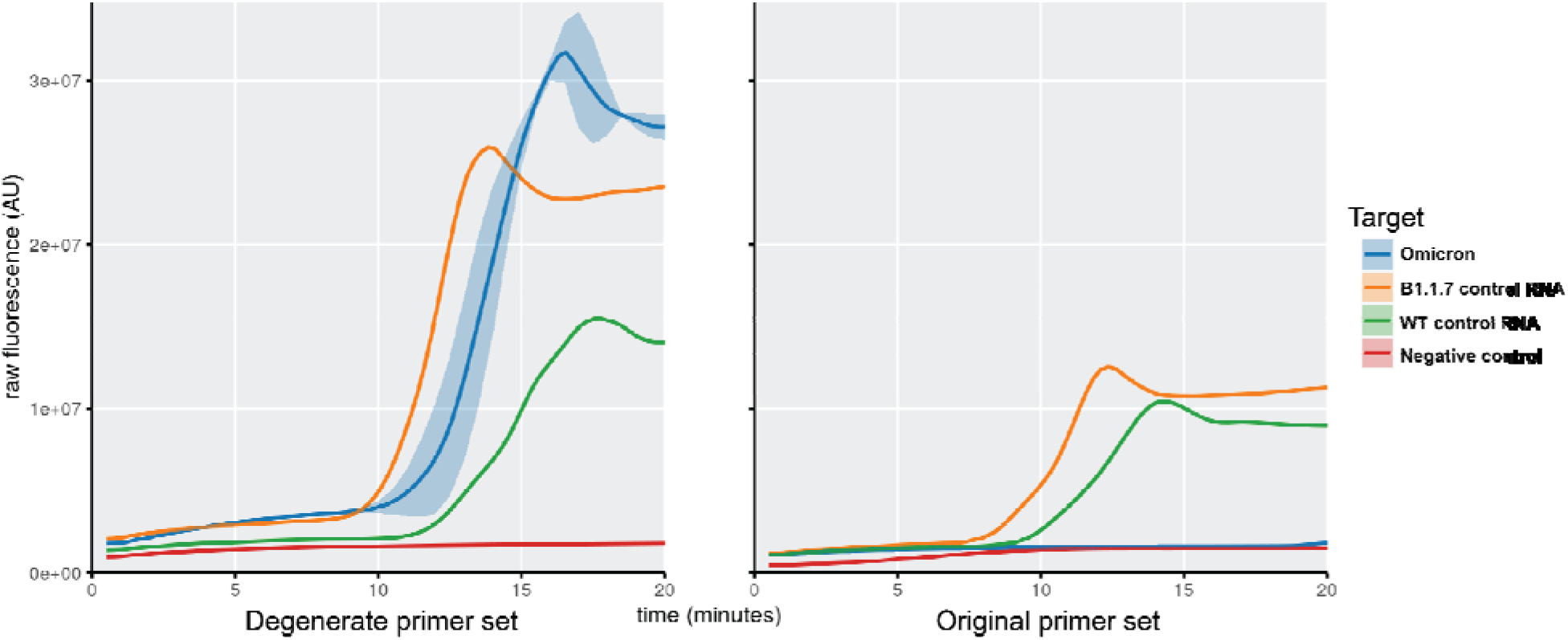
Comparison of RT-LAMP primers for processing Omicron clinical samples. Omicron clinical samples (blue), WT (green) control RNA and Alpha (orange) control RNAs were amplified using the original RT-LAMP primer set and degenerate RT-LAMP primer set. The degenerate primers amplified both the controls (WT and Alpha) and Omicron samples, whereas the original primer set only amplified the control samples (WT and Alpha).

**Extended Data Fig. 10.**
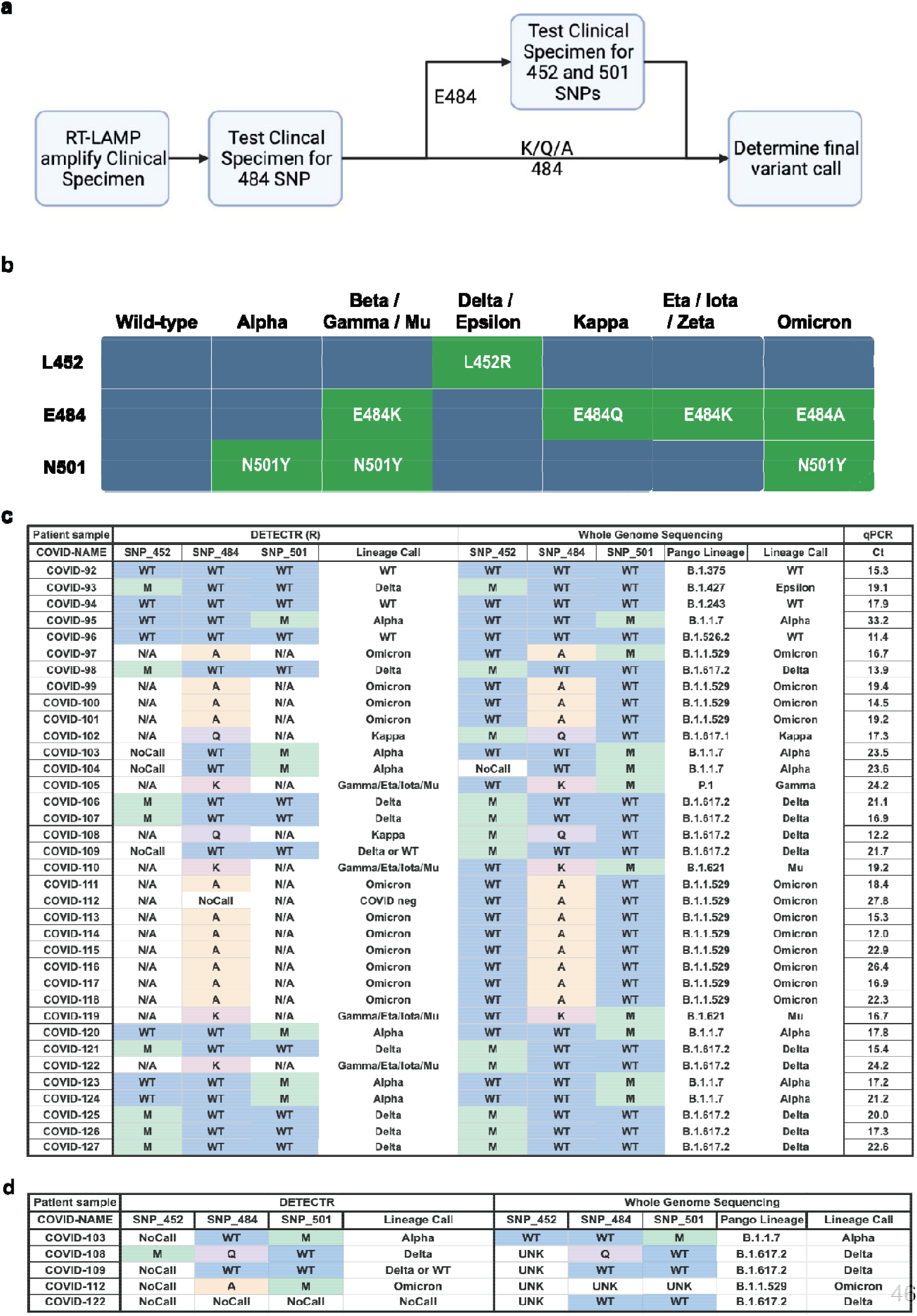
Results summary of final SNP calls by the COVID Variant DETECTR^®^ and WGS assays. **a**, Schematic of the workflow for determining the final variant calls. If the result was an A484, K484 or Q484 the final variant call was made. If the result was an E484, the sample was reflexed to DETECTR^®^ analysis at the 452 and 501 positions to make the variant determination. **b**, Interpretation table including the specific 484 SNPs. **c**, A summary table of the final SNP calls from the DETECTR^®^ assay and the SARS-CoV-2 whole genome sequencing assay including the lineage classification from DETECTR^®^ calls as well as the PANGO lineage and WHO labels assigned to the WGS calls. Ct values were obtained from the FDA EUA authorized S TaqpathTM COVID-19 RT-PCR kit. **d**, A summary table of the five discordant samples from the DETECTR^®^ assay and WGS after retesting. (NoCall = lack of data generated, N/A = assay not run)

**Supplementary Table 1.** Overall results summary of final SNP calls by the DETECTR^®^ assay and viral WGS. A summary table of the final SNP calls from the DETECTR^®^ assay and the SARS-CoV-2 whole genome sequencing assay after discordant testing. The table includes the lineage classification from DETECTR^®^ calls as well as the PANGO lineage and WHO labels assigned to the WGS calls. Ct values from running an FDA EUA authorized SARS-CoV-2 RT-PCR assay, the Taqpath™ COVID-19 RT-PCR kit, are shown. Discordant samples were reflexed back for reprocessing (*); COVID-63 was classified as a Delta variant by WGS despite its Q484 SNP call. (†).

| Patient sample | DETECTR (R) |  |  |  | Whole Genome Sequencing |  |  |  |  | qPCR |
| --- | --- | --- | --- | --- | --- | --- | --- | --- | --- | --- |
| Name | SNP 452 | SNP 484 | SNP 501 | Generic Lineage Classification | SNP 452 | SNP 484 | SNP 501 | Pango Lineage | Lineage/Variant Call | Ct |
| COVID-01 | WT | WT | WT | SARS-CoV-2 | WT | WT | WT | B.1.1.519 | N/A | 16.2 |
| COVID-02 | WT | WT | WT | SARS-CoV-2 | WT | WT | WT | B.1.1.222 | N/A | 17.2 |
| * COVID-03 | WT | WT | WT | SARS-CoV-2 | WT | WT | WT | B.1.243 | N/A | 13.9 |
| COVID-04 | M | WT | WT | Delta/Epsilon/Kappa | M | WT | WT | B.1.427 | Epsilon | 15.9 |
| COVID-05 | M | WT | WT | Delta/Epsilon/Kappa | M | WT | WT | B.1.427 | Epsilon | 22.6 |
| COVID-06 | M | WT | WT | Delta/Epsilon/Kappa | M | WT | WT | B.1.429 | Epsilon | 15.7 |
| COVID-07 | M | WT | WT | Delta/Epsilon/Kappa | M | WT | WT | B.1.427 | Epsilon | 16.1 |
| COVID-08 | WT | WT | WT | SARS-CoV-2 | WT | WT | WT | B.1.309 | N/A | 19.6 |
| COVID-09 | WT | WT | WT | SARS-CoV-2 | WT | WT | WT | B.1.1.348 | N/A | 17.2 |
| COVID-10 | M | WT | WT | Delta/Epsilon/Kappa | M | WT | WT | B.1.429 | Epsilon | 13.1 |
| COVID-11 | WT | WT | M | Alpha | WT | WT | M | B.1.1.7 | Alpha | 19.3 |
| * COVID-12 | WT | M | WT | Eta/Iota/Zeta | WT | M | WT | B.1.526 | Iota | 21.0 |
| * COVID-13 | M | WT | WT | Delta/Epsilon/Kappa | M | Q | WT | B.1.617.1 | Kappa | 17.3 |
| COVID-14 | WT | WT | M | Alpha | WT | WT | M | B.1.1.7 | Alpha | 14.9 |
| COVID-15 | WT | WT | WT | SARS-CoV-2 | WT | WT | WT | B.1.575 | N/A | 18.4 |
| COVID-16 | WT | WT | M | Alpha | WT | WT | M | B.1.1.7 | Alpha | 21.6 |
| COVID-17 | WT | WT | M | Alpha | WT | WT | M | B.1.1.7 | Alpha | 13.2 |
| COVID-18 | M | WT | WT | Delta/Epsilon/Kappa | M | WT | WT | B.1.280 | N/A | 13.9 |
| COVID-19 | M | WT | WT | Delta/Epsilon/Kappa | M | WT | WT | B.1.617.2 | Delta | 18.9 |
| * COVID-20 | WT | WT | M | Alpha | WT | WT | M | B.1.1.7 | Alpha | 23.5 |
| COVID-21 | WT | WT | M | Alpha | WT | WT | M | B.1.1.7 | Alpha | 25.9 |
| COVID-22 | WT | WT | M | Alpha | WT | WT | M | B.1.1.7 | Alpha | 22.2 |
| COVID-23 | WT | M | M | Beta/Gamma/Mu | WT | M | M | P1 | Gamma | 17.1 |
| COVID-24 | WT | WT | M | Alpha | WT | WT | M | B.1.1.7 | Alpha | 23.6 |
| COVID-25 | WT | M | M | Beta/Gamma/Mu | WT | M | M | P1 | Gamma | 20.4 |
| COVID-26 | WT | M | M | Beta/Gamma/Mu | WT | M | M | P1 | Gamma | 21.4 |
| COVID-27 | WT | M | M | Beta/Gamma/Mu | WT | M | M | P1 | Gamma | 25.0 |
| COVID-28 | WT | M | M | Beta/Gamma/Mu | WT | M | M | P1 | Gamma | 15.2 |
| COVID-29 | WT | WT | M | Alpha | WT | WT | M | B.1.1.7 | Alpha | 16.6 |
| COVID-30 | WT | M | M | Beta/Gamma/Mu | WT | M | M | P1 | Gamma | 17.5 |
| * COVID-31 | WT | WT | M | Alpha | NoCall | WT | M | B.1.1.7 | Alpha | 23.6 |
| COVID-32 | WT | M | WT | Eta/Iota/Zeta | WT | M | WT | B.1.1.318 | N/A | 15.6 |
| COVID-33 | WT | WT | M | Alpha | WT | WT | M | B.1.1.7 | Alpha | 22.0 |
| COVID-34 | WT | M | M | Beta/Gamma/Mu | WT | M | M | P1 | Gamma | 17.2 |
| COVID-35 | WT | M | M | Beta/Gamma/Mu | WT | M | M | P1 | Gamma | 18.9 |
| COVID-36 | WT | M | M | Beta/Gamma/Mu | WT | M | M | P1 | Gamma | 18.9 |
| COVID-37 | WT | M | M | Beta/Gamma/Mu | WT | M | M | P1 | Gamma | 18.5 |
| COVID-38 | WT | M | M | Beta/Gamma/Mu | WT | M | M | P1 | Gamma | 15.4 |
| COVID-39 | WT | M | M | Beta/Gamma/Mu | WT | M | M | P1 | Gamma | 16.4 |
| COVID-40 | WT | M | M | Beta/Gamma/Mu | WT | M | M | P1 | Gamma | 16.7 |
| * COVID-41 | WT | M | M | Beta/Gamma/Mu | WT | M | M | P1 | Gamma | 24.2 |
| COVID-42 | WT | M | M | Beta/Gamma/Mu | WT | M | M | P1 | Gamma | 17.7 |
| COVID-43 | WT | WT | M | Alpha | WT | WT | M | B.1.1.7 | Alpha | 15.3 |
| COVID-44 | WT | WT | M | Alpha | WT | WT | M | B.1.1.7 | Alpha | 17.9 |
| COVID-45 | WT | M | M | Beta/Gamma/Mu | WT | M | M | P1 | Gamma | 16.6 |
| COVID-46 | M | WT | WT | Delta/Epsilon/Kappa | M | WT | WT | B.1.617.2 | Delta | 23.2 |
| COVID-47 | M | WT | WT | Delta/Epsilon/Kappa | M | WT | WT | B.1.526.1 | N/A | 16.6 |
| COVID-48 | WT | WT | M | Alpha | WT | WT | M | B.1.1.7 | Alpha | 13.7 |
| COVID-49 | M | WT | WT | Delta/Epsilon/Kappa | M | WT | WT | B.1.617.2 | Delta | 17.9 |
| COVID-50 | WT | WT | M | Alpha | WT | WT | M | B.1.1.7 | Alpha | 15.8 |
| COVID-51 | WT | WT | M | Alpha | WT | WT | M | B.1.1.7 | Alpha | 15.8 |
| COVID-52 | M | WT | WT | Delta/Epsilon/Kappa | M | WT | WT | AY.1 | Delta | 18.1 |
| COVID-53 | M | WT | WT | Delta/Epsilon/Kappa | M | WT | WT | AY.1 | Delta | 22.9 |
| COVID-54 | M | WT | WT | Delta/Epsilon/Kappa | M | WT | WT | AY.1 | Delta | 22.0 |
| COVID-55 | M | WT | WT | Delta/Epsilon/Kappa | M | WT | WT | B.1.617.2 | Delta | 19.4 |
| * COVID-56 | M | WT | WT | Delta/Epsilon/Kappa | M | WT | WT | B.1.617.2 | Delta | 21.1 |
| COVID-57 | M | WT | WT | Delta/Epsilon/Kappa | M | WT | WT | B.1.526.1 | N/A | 26.6 |
| COVID-58 | M | WT | WT | Delta/Epsilon/Kappa | M | WT | WT | B.1.617.2 | Delta | 17.8 |
| COVID-59 | M | WT | WT | Delta/Epsilon/Kappa | M | WT | WT | AY.1 | Delta | 18.5 |
| COVID-60 | M | WT | WT | Delta/Epsilon/Kappa | M | WT | WT | AY.1 | Delta | 21.5 |
| * COVID-61 | M | WT | WT | Delta/Epsilon/Kappa | M | WT | WT | B.1.617.2 | Delta | 16.9 |
| COVID-62 | M | WT | WT | Delta/Epsilon/Kappa | M | WT | WT | B.1.617.2 | Delta | 14.1 |
| * COVID-63 | M | WT | WT | Delta/Epsilon/Kappa | M | Q | WT | B.1.617.2 | Delta | 12.2 |
| COVID-64 | M | WT | WT | Delta/Epsilon/Kappa | M | WT | WT | B.1.617.2 | Delta | 22.0 |
| COVID-65 | M | WT | WT | Delta/Epsilon/Kappa | M | WT | WT | B.1.617.2 | Delta | 21.9 |
| COVID-66 | M | WT | WT | Delta/Epsilon/Kappa | M | WT | WT | B.1.617.2 | Delta | 14.8 |
| COVID-67 | M | WT | WT | Delta/Epsilon/Kappa | M | WT | WT | B.1.617.2 | Delta | 18.7 |
| COVID-68 | M | WT | WT | Delta/Epsilon/Kappa | M | WT | WT | B.1.617.2 | Delta | 16.6 |
| COVID-69 | M | WT | WT | Delta/Epsilon/Kappa | M | WT | WT | B.1.617.2 | Delta | 19.0 |
| COVID-70 | M | WT | WT | Delta/Epsilon/Kappa | M | WT | WT | B.1.617.2 | Delta | 21.1 |
| COVID-71 | M | WT | WT | Delta/Epsilon/Kappa | M | WT | WT | B.1.617.2 | Delta | 14.0 |
| COVID-72 | M | WT | WT | Delta/Epsilon/Kappa | M | WT | WT | B.1.617.2 | Delta | 25.6 |
| * COVID-73 | M | WT | WT | Delta/Epsilon/Kappa | M | WT | WT | B.1.617.2 | Delta | 21.7 |
| COVID-74 | WT | M | M | Beta/Gamma/Mu | WT | M | M | B.1.623 | N/A | 23.7 |
| COVID-75 | M | WT | WT | Delta/Epsilon/Kappa | M | WT | WT | B.1.617.2 | Delta | 17.7 |
| COVID-76 | M | WT | WT | Delta/Epsilon/Kappa | M | WT | WT | AY.1 | Delta | 20.5 |
| COVID-77 | M | WT | WT | Delta/Epsilon/Kappa | M | WT | WT | B.1.617.2 | Delta | 25.2 |
| COVID-78 | M | WT | WT | Delta/Epsilon/Kappa | M | WT | WT | B.1.617.2 | Delta | 18.7 |
| COVID-79 | M | WT | WT | Delta/Epsilon/Kappa | M | WT | WT | B.1.617.2 | Delta | 19.3 |
| COVID-80 | M | WT | WT | Delta/Epsilon/Kappa | M | WT | WT | B.1.617.2 | Delta | 18.4 |
| * COVID-81 | WT | M | M | Beta/Gamma/Mu | WT | M | M | B.1.621 | Mu | 19.2 |
| COVID-82 | M | WT | WT | Delta/Epsilon/Kappa | M | WT | WT | AY.1 | Delta | 18.3 |
| COVID-83 | M | WT | WT | Delta/Epsilon/Kappa | M | WT | WT | A.2.5 | Delta | 17.6 |
| COVID-84 | M | WT | WT | Delta/Epsilon/Kappa | M | WT | WT | AY.1 | Delta | 12.7 |
| COVID-85 | M | WT | WT | Delta/Epsilon/Kappa | M | WT | WT | B.1.617.2 | Delta | 21.0 |
| COVID-86 | M | WT | WT | Delta/Epsilon/Kappa | M | WT | WT | B.1.617.2 | Delta | 16.8 |
| COVID-87 | M | WT | WT | Delta/Epsilon/Kappa | M | WT | WT | B.1.617.2 | Delta | 17.7 |
| COVID-88 | WT | M | WT | Eta/Iota/Zeta | WT | M | WT | B.1.526 | Iota | 14.8 |
| COVID-89 | M | WT | WT | Delta/Epsilon/Kappa | M | WT | WT | B.1.617.2 | Delta | 20.5 |
| COVID-90 | M | WT | WT | Delta/Epsilon/Kappa | M | WT | WT | B.1.617.2 | Delta | 21.6 |
| COVID-91 | M | WT | WT | Delta/Epsilon/Kappa | M | WT | WT | A.2.5 | Delta | 15.6 |

**Supplementary Table 2.** Summary of samples showing no COVID detection. A summary table of the clinical specimens with no signal in the RT-LAMP nor the DETECTR^®^ reactions. These samples were called “COVID Not Detected (ND)” by both the COVID-19 variant DETECTR^®^ assay and the SARS-CoV-2 whole genome sequencing assays. Ct values were obtained from the FDA EUA authorized S Taqpath^TM^ COVID-19 RT-PCR kit. (NoCall = lack of data generated, N/A = assay not run).

| Patient specimen | DETECTR (R) |  |  |  | Whole Genome Sequencing |  |  |  |  | qPCR |
| --- | --- | --- | --- | --- | --- | --- | --- | --- | --- | --- |
| Sample Name | SNP_452 | SNP_484 | SNP_501 | Lineage Call | SNP_452 | SNP_484 | SNP_501 | Pango Lineage | Lineage/Variant Call | Ct |
| Sample-1 | N/A | NoCall | N/A | COVID ND | NoCall | NoCall | NoCall | COVID neg | COVID ND | N/A |
| Sample-2 | N/A | NoCall | N/A | COVID ND | NoCall | NoCall | NoCall | COVID neg | COVID ND | N/A |
| Sample-3 | N/A | NoCall | N/A | COVID ND | NoCall | NoCall | NoCall | > 30 Ct | COVID ND | 38.6 |
| Sample-4 | N/A | NoCall | N/A | COVID ND | NoCall | NoCall | NoCall | > 30 Ct | COVID ND | 33.2 |
| Sample-5 | N/A | NoCall | N/A | COVID ND | NoCall | NoCall | NoCall | COVID neg | COVID ND | N/A |
| Sample-6 | N/A | NoCall | N/A | COVID ND | NoCall | NoCall | NoCall | > 30 Ct | COVID ND | 38.7 |
| Sample-7 | N/A | NoCall | N/A | COVID ND | NoCall | NoCall | NoCall | > 30 Ct | COVID ND | 35.8 |
| Sample-8 | N/A | NoCall | N/A | COVID ND | NoCall | NoCall | NoCall | > 30 Ct | COVID ND | 32.1 |
| Sample-9 | N/A | NoCall | N/A | COVID ND | NoCall | NoCall | NoCall | > 30 Ct | COVID ND | 30.1 |
| Sample-10 | N/A | NoCall | N/A | COVID ND | NoCall | NoCall | NoCall | > 30 Ct | COVID ND | 36.9 |
| Sample-11 | N/A | NoCall | N/A | COVID ND | NoCall | NoCall | NoCall | > 30 Ct | COVID ND | 34.7 |
| Sample-12 | N/A | NoCall | N/A | COVID ND | NoCall | NoCall | NoCall | Covid neg | COVID ND | N/A |

**Supplementary Table 3.**
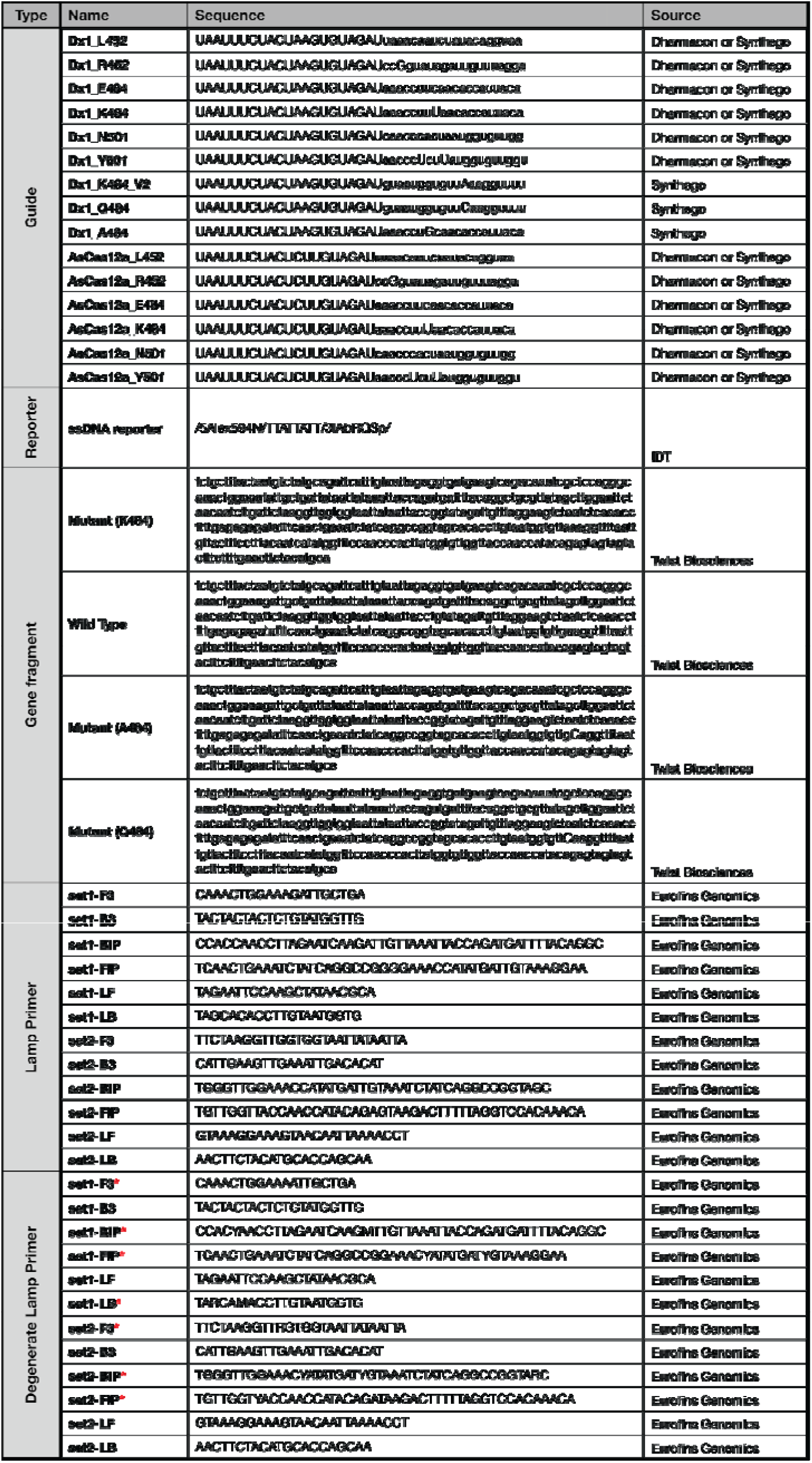
Nucleic acid sequences used in this study. A list of guide RNAs, reporter molecules, LAMP primers and synthetic gene fragment targets with their respective suppliers. (* indicates LAMP primers that contain degenerate nucleotides)

